# Impaired resolution of blood transcriptomes through tuberculosis treatment with diabetes comorbidity

**DOI:** 10.1101/2022.02.07.22269422

**Authors:** Clare Eckold, Cassandra L.R. van Doorn, Rovina Ruslami, Katharina Ronacher, Anca-Lelia Riza, Suzanne van Veen, Ji-Sook Lee, Vinod Kumar, Sarah Kerry-Barnard, Stephanus T. Malherbe, Léanie Kleynhans, Kim Stanley, Simone A. Joosten, Julia A Critchley, Philip C. Hill, Reinout van Crevel, Cisca Wijmenga, Mariëlle C. Haks, Mihai Ioana, Bachti Alisjahbana, Gerhard Walzl, Tom H. M. Ottenhoff, Hazel M. Dockrell, Eleonora Vianello, Jacqueline M. Cliff, the TANDEM Consortium

## Abstract

**Background:** People with diabetes are more likely to develop tuberculosis (TB) and to have poor TB treatment outcomes than those without. We previously showed that blood transcriptomes in people with TB-diabetes (TB-DM) co-morbidity have excessive inflammatory and reduced interferon responses at TB diagnosis. It is unknown whether this persists through treatment, potentially underlying adverse outcomes.

**Methods:** Pulmonary TB patients were recruited in South Africa, Indonesia and Romania, and classified as having TB-DM, TB with prediabetes, TB-related hyperglycaemia or uncomplicated TB, based on glycated haemoglobin (HbA1c) concentration at TB diagnosis and after 6 months of TB treatment. Gene expression in blood samples collected at diagnosis and at regular intervals throughout treatment was measured by unbiased RNA-Seq and targeted Multiplex Ligation-dependent Probe Amplification.

**Results:** Gene expression was modulated by TB treatment in all groups but to different extents, such that differences remained in people with TB-DM relative to TB-only throughout, including genes involved in innate responses, anti-microbial immunity and the inflammasome. People with prediabetes or with TB-related hyperglycaemia had gene expression more similar to people with TB-DM than TB-only throughout treatment. The overall pattern of change was similar across clinical groups irrespective of glycaemic index, permitting models predictive of TB treatment to be developed.

**Conclusions:** The exacerbated transcriptome changes seen in TB-DM take longer to resolve during TB treatment, indicating that prolonged treatment or host-directed therapy may be needed to improve TB treatment outcomes. Development of transcriptome-based biomarker signatures of TB-treatment response should include people with diabetes to be useful across populations.

**Key Points:** Host blood transcriptomes are altered in tuberculosis, and further altered with diabetes co-morbidity. We have shown that there is similar resolution of transcriptomes through treatment, but with differing magnitude and kinetics in TB patients with or without diabetes.

## Introduction

Diabetes mellitus (DM) negatively impacts TB control by increasing the risk of *Mycobacterium tuberculosis* infection [1] and of progression to active TB disease three-fold [2, 3]. The growing prevalence of DM, particularly in countries with high TB burden, means DM now underlies around 15% of TB cases globally [4], accounting for 10% of TB deaths in HIV-negative people. Concomitant DM negatively impacts TB-treatment outcomes, with increased risk of delayed sputum conversion, relapse, treatment failure and death: relative risk for each poor outcome is ∼2 to ∼5 in meta-analyses [5, 6]. It is unknown whether extending standard TB treatment would improve outcome for TB-DM comorbid patients, or whether alternative treatment is required, such as host-directed therapy.

Worldwide DM prevalence is ∼463 million people, estimated to rise to 700 million by 2045 [7]. The majority of people have type-2 DM, caused by a reduction in insulin’s ability to control target cell metabolism triggering an increase in insulin production, pancreatic damage through exhaustion, and impaired glucose tolerance. There is a spectrum from normal through to full DM via intermediate hyperglycaemia (IH). People with IH are more likely to develop DM in the future [8]. HbA1c concentration can indicate an individual’s position on this spectrum [8]. Infectious diseases, including TB, can cause temporary stress hyperglycaemia, which carries a higher risk of adverse events than longer-term pre-diabetes [9]. TB-induced stress hyperglycaemia also makes DM diagnosis difficult: some people with apparent newly-diagnosed DM at TB diagnosis no longer reach DM diagnostic criteria after TB treatment [10]. TB incidence and TB-DM treatment outcomes are worse in people with poorly controlled DM with higher HbA1c concentrations [11].

People with TB-DM comorbidity have altered immunity compared to uncomplicated TB, with both innate and adaptive immune responses affected [12]. In plasma, various inflammatory cytokines such as IL-1β, IL-17A, IFNγ and TNFα are more elevated in TB-DM [13, 14] and TB-pre-diabetes [15] than in uncomplicated TB. People with TB-DM have more circulating Th1 and Th17 cells and fewer Tregs. In uncomplicated TB, peripheral immune responses typically resolve to normal levels during successful TB treatment [14]. In contrast, the excessive inflammatory plasma cytokine responses in TB-DM are still evident after treatment completion [16], and dendritic cell, monocyte [17] and T cell differentiation [18] aberrations are still present at 2 months, although resolved by 6 months, indicating a delayed response to TB treatment in TB-DM patients.

Transcriptomic technologies have delineated altered peripheral immunity in TB in multiple studies, revealing an enhanced circulating inflammatory and type 1 interferon response [19-22]. With successful TB treatment, the transcriptomic signature is rapidly downregulated, has largely diminished after two months of treatment and mostly disappears by 12 months [20, 22, 23], mirroring clinical resolution and chest X-ray improvement; however transcriptomes do not fully resolve with poor TB treatment outcome [24]. We recently showed [25] that DM comorbidity, as well as IH, significantly affects the TB diagnosis biosignature, causing an enhanced inflammatory but reduced interferon response. The impact of DM on transcriptomes through TB treatment has not been described. The aim of this study was to determine whether transcriptomic biosignatures resolve normally in TB-DM, or whether changes during TB treatment are kinetically or qualitatively different to those in uncomplicated TB.

## Methods

### Patient recruitment and classification

Newly diagnosed patients with bacteriologically confirmed pulmonary TB, with or without concomitant DM, were recruited in three locations: Bandung, Indonesia (UNPAD), Cape Town, South Africa (SUN) and Craiova, Romania (UMFCV), as part of the TANDEM project [26]. Exclusion criteria were multi-drug-resistant TB, HIV positivity, pregnancy, other serious co-morbidity or corticosteroid use. All participants gave written informed consent.

The study was approved by LSHTM Observational Research Ethics Committee (6449/July2013), SUN Health Research Ethics Committee (N13/05/064/July2013), UNPAD Health Research Ethics Committee (377/UN6.C2.1.2/ KEPK/PN/2012), and UMFCV Committee of Ethics and Academic and Scientific Deontology (94/September2013).

All TB patients underwent first line TB treatment according to WHO guidelines. Most patients diagnosed with DM received local standard of care treatment, and medication taken was noted. A TB-DM subgroup within the Indonesian cohort had intensive HbA1c monitoring as part of a pragmatic randomised control trial, with DM medication changed accordingly [27]. Participants were classified by DM/glycaemia status at TB diagnosis and after 6 months TB treatment (Supplementary Table S1). The “TB-DM” group included patients with both pre-existing and newly diagnosed DM (Supplementary Table S2) Newly diagnosed TB-DM had laboratory HbA1c test ≥6.5% with confirmatory HbA1c test ≥6.5% or fasting blood glucose ≥7 mmol/L at TB diagnosis [26, 28], followed by a further HbA1c test ≥6.5% after 6 months of TB treatment. Patients whose HbA1c test results were ≥5.7% and <6.5% at both TB diagnosis and at 6 months were deemed to have pre-diabetes (“TB-preDM”). Patients whose HbA1c result was ≥5.7% at TB diagnosis but below <5.7% at 6 months were deemed to have TB-related intermediate hyperglycaemia at TB diagnosis (“TBrel-IH”). All groups were evenly sex balanced, except for male predominance in TB-PreDM. Age ranges were similar across groups. In the Indonesian TB-DM group, there was a highly significant decrease in HbA1c through TB treatment, likely due to intensive DM follow-up (Supplementary Figure S1 & Supplementary Table S1); this was not evident in South Africa or Romania.

### Sample collection and RNA extraction

Venous blood samples (2.5ml) were collected into PAXgene Blood RNA Tubes (PreAnalytiX) from TB patients prior to TB treatment initiation (W0) and at intervals through treatment (W2,4,8,16,26) up to 12 months post diagnosis (W52), and stored at -80^°^C prior to analysis. Total RNA was extracted using RNeasy spin columns (Qiagen) and quantified by Nanodrop (Agilent).

### Unbiased whole genome RNA-Seq

A detailed description of RNA-Seq analysis is given in Supplementary Methods. Samples were processed using the poly-A tail Bioscientific NEXTflex-Rapid-Directional mRNA-seq method and single-end sequenced. Longitudinal differential gene expression analyses were performed on normalised data using the MaSigPro (v1.62.0) [29] package in R. MaSigPro follows a two-step regression method to find genes with significant temporal expression changes and significant differences between groups. Modular analysis was performed on genes that were differentially expressed between clinical groups.

### Targeted gene expression profiling

Targeted gene expression profiling was performed using dual-color Reverse-Transcriptase Multiplex Ligation-dependent Probe Amplification (dcRT-MLPA) [30]: a detailed description is given in Supplementary Methods. Primers and half-probes were designed for 4 housekeeping genes and 144 selected key immune TB-related genes (Supplementary Table S3). Longitudinal differentially expressed genes (DEGs) within groups were identified using linear mixed models. Signatures with the best discriminatory capability were identified using logistic regression with lasso regularization (glmnet R package).

## Results

### Global longitudinal transcriptomes in TB-DM

Gene expression was determined in venous blood by RNA-Seq in a subgroup of study participants from the four TB patient clinical groups (Supplementary Table S1; Supplementary Figure S1). The molecular degree of perturbation (MDP) of gene expression in individual samples from patients with TB-only or TB-DM over time was calculated relative to mean gene expression at diagnosis in TB-only (Figure 1). As expected, there were gene expression changes during TB treatment in the TB-only group, evident by week 2 and continuing throughout treatment. Global gene expression was perturbed in patients with TB-DM relative to TB-only at TB diagnosis, and while there was some resolution through time, the changes were reduced such that global gene expression in the TB-DM group remained different to the TB-only group at all time points (Figure 1).

**Figure 1:**
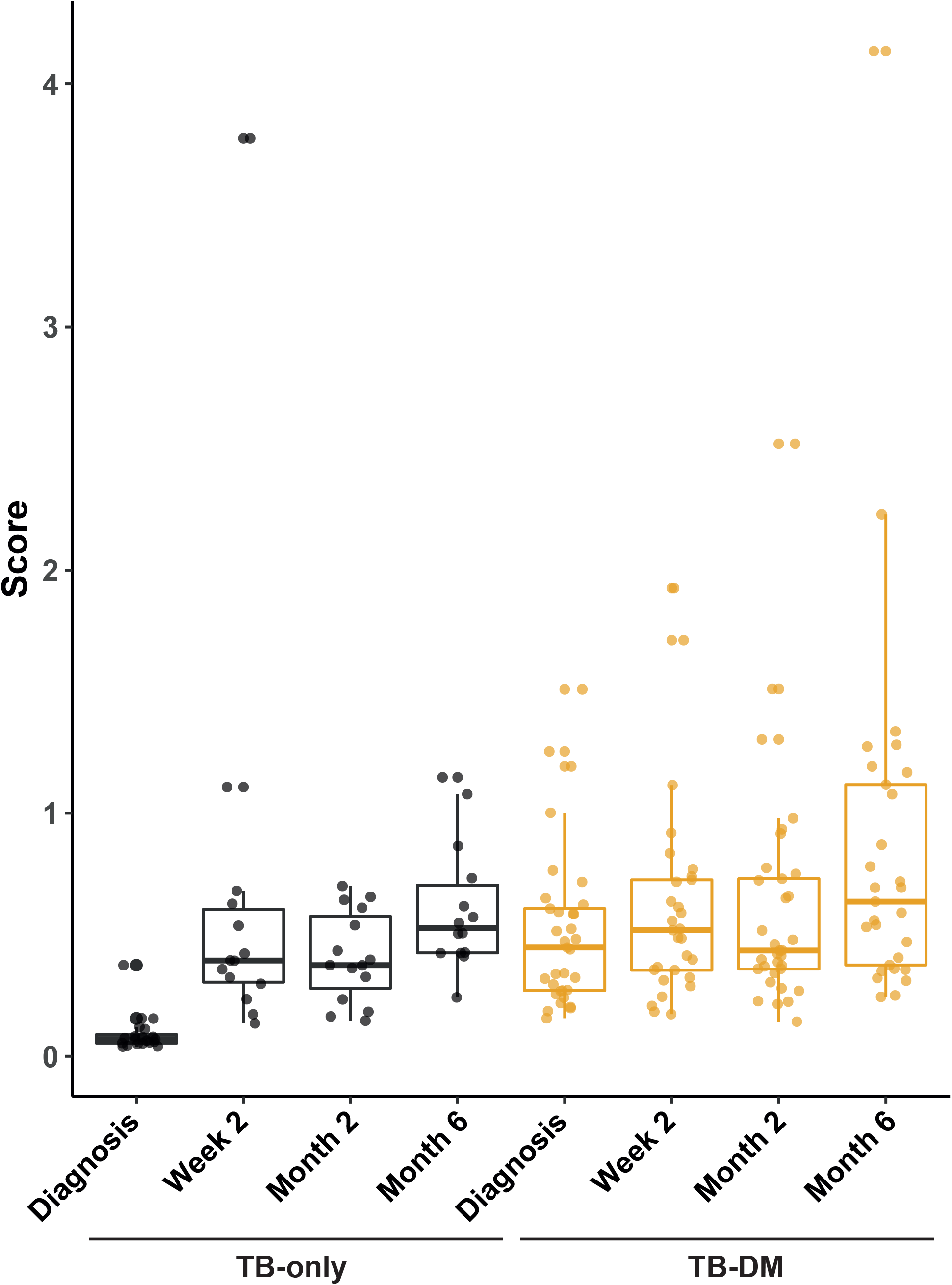
Molecular Degree of Perturbation Plots representing change in global gene expression in blood relative to patients with TB-only at TB diagnosis. Gene expression was determined by RNA-Seq of whole venous blood from pulmonary TB patients from all three clinical locations with (TB-DM: n=34) or without (TB-only: n= 18) concomitant diabetes, at TB diagnosis and during TB treatment. The bars show the median and 1.5*inter-quartile range.

The MaSigPro analysis package identified 167 genes with significantly different changed expression between TB-DM and TB-only groups through TB treatment, in the combined dataset from South Africa, Indonesia and Romania. Hierarchical clustering of these genes based on similar expression patterns yielded 9 clusters (Figure 2; Supplementary Table S4). Clusters which were more highly expressed in TB-DM patients throughout treatment (clusters 1,2,4&8) were enriched for genes involved in the innate immune response, IL-4 signalling, protein dimerisation and neutrophil chemotaxis, determined using the DAVID Functional Annotation Tool [31] (Table 1). Cluster 6 exhibited divergence between TB and TB-DM patients only at week 8 of treatment: this cluster was enriched for genes involved in anti-viral and IFN signalling responses. Clusters more highly expressed in TB-only patients (clusters 5,7,9) were smaller and enriched for alternative splice variants and immunoglobulins.

**Figure 2:**
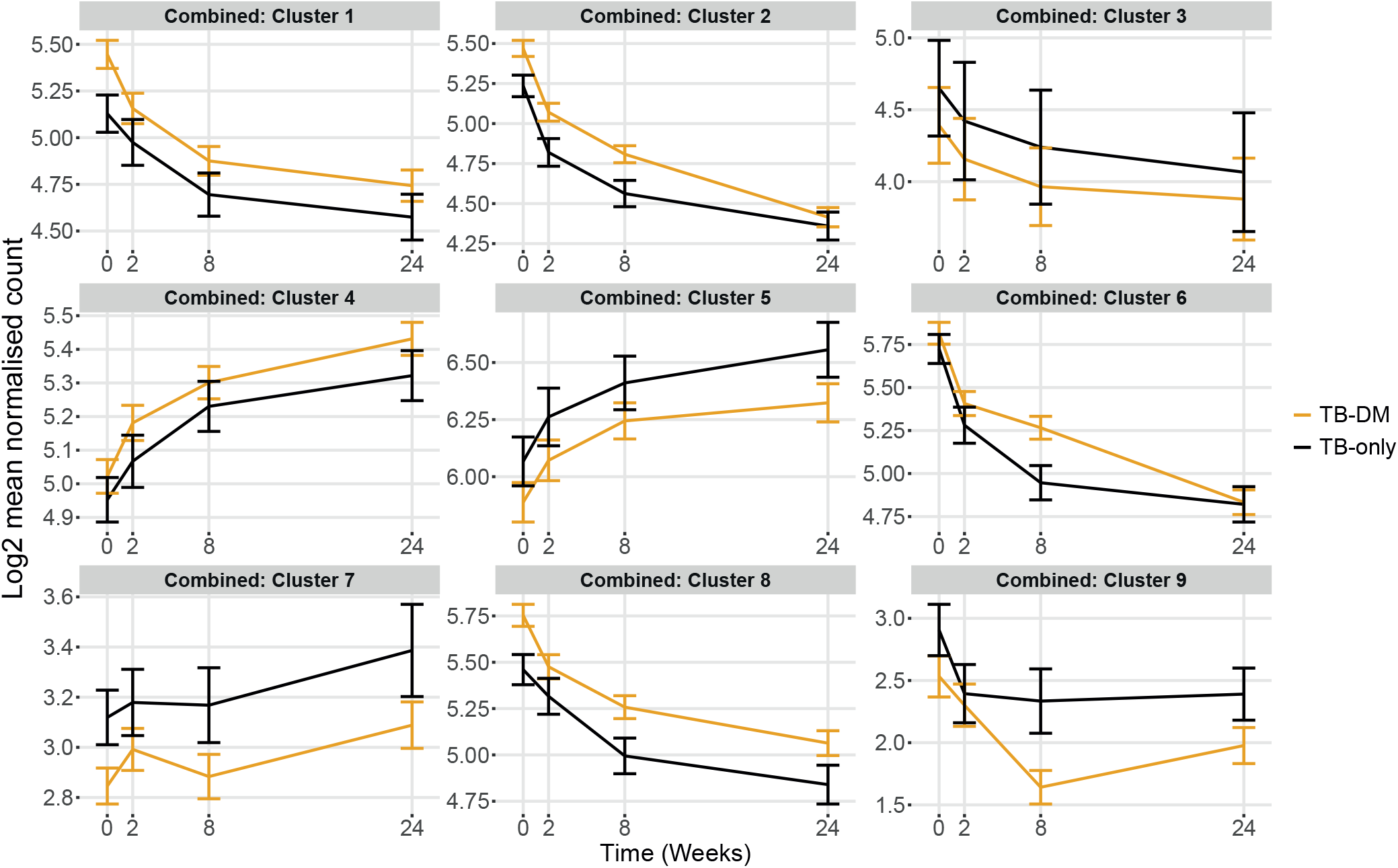
MaSigPro analysis of change in gene expression through TB treatment in blood samples from patients in all 3 populations combined (South Africa, Indonesia and Romania). MaSigPro identified genes that behave similarly between patient groups using hierarchical clustering. Results are shown for log-transformed normalised count for the TB-only group or TB-DM. Bars show mean ± 1 SEM. Data were filtered to remove lowly abundant transcripts prior to analysis.

**Table 1:** Clusters of genes differentially expressed between TB-DM and TB-only patients in MaSigPro analysis of the combined RNA-Seq dataset from South Africa, Indonesia and Romania.

| Cluster | Overall pattern | Number of transcripts | Gene Function |  |  |  | Top Non-redundant Functions from DAVID<sup>#</sup> |
| --- | --- | --- | --- | --- | --- | --- | --- |
|  |  |  | Protein Coding | Processed Transcript | Pseudo-gene | Regulatory RNAs<sup>*</sup> |  |
| 1 | Higher in TB-DM throughout; decreasing with time | 27 | 19 | 0 | 2 | 6 | Innate Immunity<br>Antimicrobial<br>RAGE receptor binding |
| 2 | Higher in TB-DM throughout; decreasing with time | 50 | 28 | 5 | 4 | 13 | IPAF inflammasome<br>IL-4 signalling<br>Transmembrane helices |
| 3 | Lower in TB-DM throughout; decreasing with time | 6 | 6 | 0 | 0 | 0 | Disulphide bond<br>Inflammation/fibrosis |
| 4 | Higher in TB-DM throughout; increasing with time | 17 | 11 | 3 | 0 | 3 | Coiled coil<br>Protein homo-dimersiation |
| 5 | Lower in TB-DM throughout; increasing with time | 9 | 8 | 1 | 0 | 0 | Collagen-binding<br>Alternative splicing<br>phosphoprotein |
| 6 | Higher in TB-DM at week 8, otherwise similar | 28 | 22 | 1 | 0 | 5 | GTPase activity<br>Anti-viral defence<br>IFNγ signalling |
| 7 | Lower in TB-DM throughout; rising end treatment | 10 | 4 | 1 | 0 | 5 | Splice variant |
| 8 | Higher in TB-DM throughout; decreasing with time | 16 | 13 | 2 | 0 | 1 | Secreted<br>Neutrophil chemotaxis<br>Transmembrane helix |
| 9 | Lower in TB-DM at week 8 and 24, otherwise similar | 4 | 3 | 0 | 0 | 1 | Immunoglobulins |

<sup>#</sup>DAVID [31] analysis of GO terms BP, MF, CC direct UP-Keywords
<sup>\*</sup>Retained introns, Antisense, LncRNA, miRNA, nonsense-mediated decay, sense overlapping, sense intronic, snoRNA,

### Aberrant longitudinal transcriptomes in TB patients with intermediate hyperglycaemia

Previously [25] we showed gene expression in TBrel-IH is more similar to people with diagnosed DM than with TB-only at TB diagnosis. We repeated the MaSigPro analyses separately for South Africa and Indonesia, combining all those patients with raised glycaemic indices but not DM at TB diagnosis, to determine how transcriptomes changed through TB treatment in intermediate groups (Supplementary Figure S2). In South Africa, the analysis resulted in 1,179 transcripts separated into three hierarchical clusters, which changed through treatment differently across clinical groups (Supplementary Figure S2A; Supplementary Table S5), with the combined intermediate group behaving more similarly to TB-DM. Similar results were obtained with the Indonesian cohort, with 2,354 transcripts across 4 hierarchical clusters behaving differently between clinical groups (Supplementary Figure S2B; Supplementary Table S6).

A core list of 102 genes overlapped between MaSigPro analyses for the combined cohort from Romania, South Africa and Indonesia, and from the latter two populations separately (Supplementary Figure S3; Supplementary Table S7). Gene ontology and pathway analyses of this core list using the g:profiler webtool revealed functional enrichment of genes involved in the immune response, response to biotic stimuli, and gene products localising to intracellular vesicles (Supplementary Figure S4). We hypothesised there would be differences between the TB-preDM and TBrel-IH groups, due to persistence or resolution of hyperglycaemia through TB treatment; however, both TB-preDM and TBrel-IH patients responded more similarly to TB-DM than to TB-only patients (Figure 3). Longitudinal mixed effects model analysis of mean expression within core gene list clusters showed highly significant changes across all four clinical groups throughout treatment, with differences between the groups in larger gene clusters (Supplementary Table S8). Importantly, there was no interaction between clinical group and time, showing there was resolution of expression in all groups through treatment, albeit from different starting points and at different rates.

**Figure 3:**
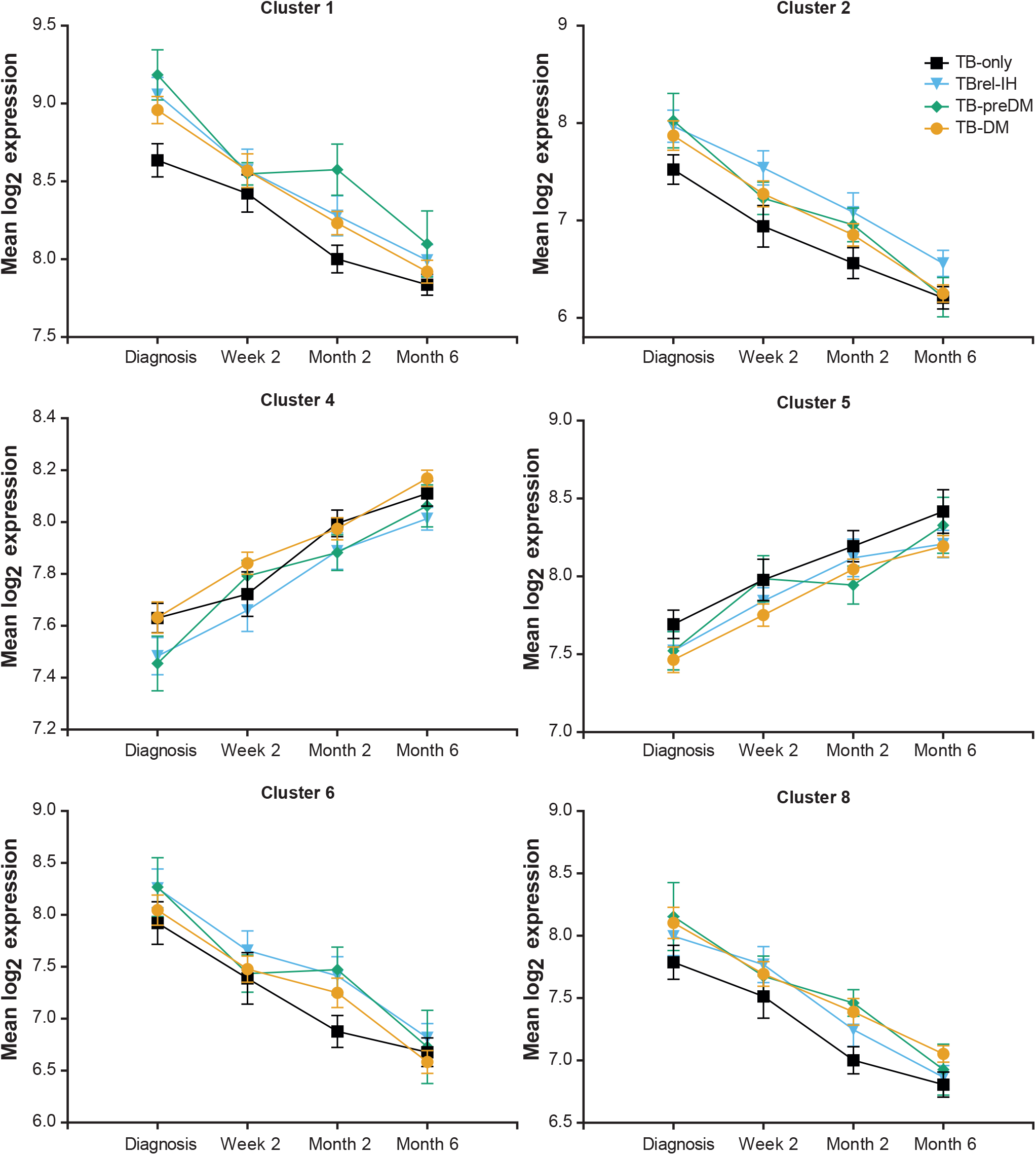
Gene expression through treatment in TB patients with Pre-diabetes or TB-related intermediate hyperglycaemia, relative to TB-DM and TB-only patients. The expression of genes in the Core 102 genelist (Supplementary Table S7) was summed for those genes within each MaSigPro gene cluster (Figure 2) for individual patients (log_2_ scale). Only MaSigPro clusters with > 3 genes in the core gene list are shown. Points show the mean ± SEM for each of the four clinical groups at each timepoint.

### Modular analysis of differentially expressed genes (DEGs)

DEGs identified in MaSigPro analyses were used in modular analyses to understand biological differences between clinical groups in South Africa and Indonesia (Supplementary Table 9 and 10 respectively). The most statistically significant modules were investigated further by calculating their modular activity in TB-DM relative to TB-only through time. The top module in both populations was immune activation, which was upregulated in TB-DM compared to TB-only throughout treatment. In both populations, different modules fluctuated between TB-DM and TB, and behaved inversely to one another through treatment (Figure 4).

**Figure 4:**
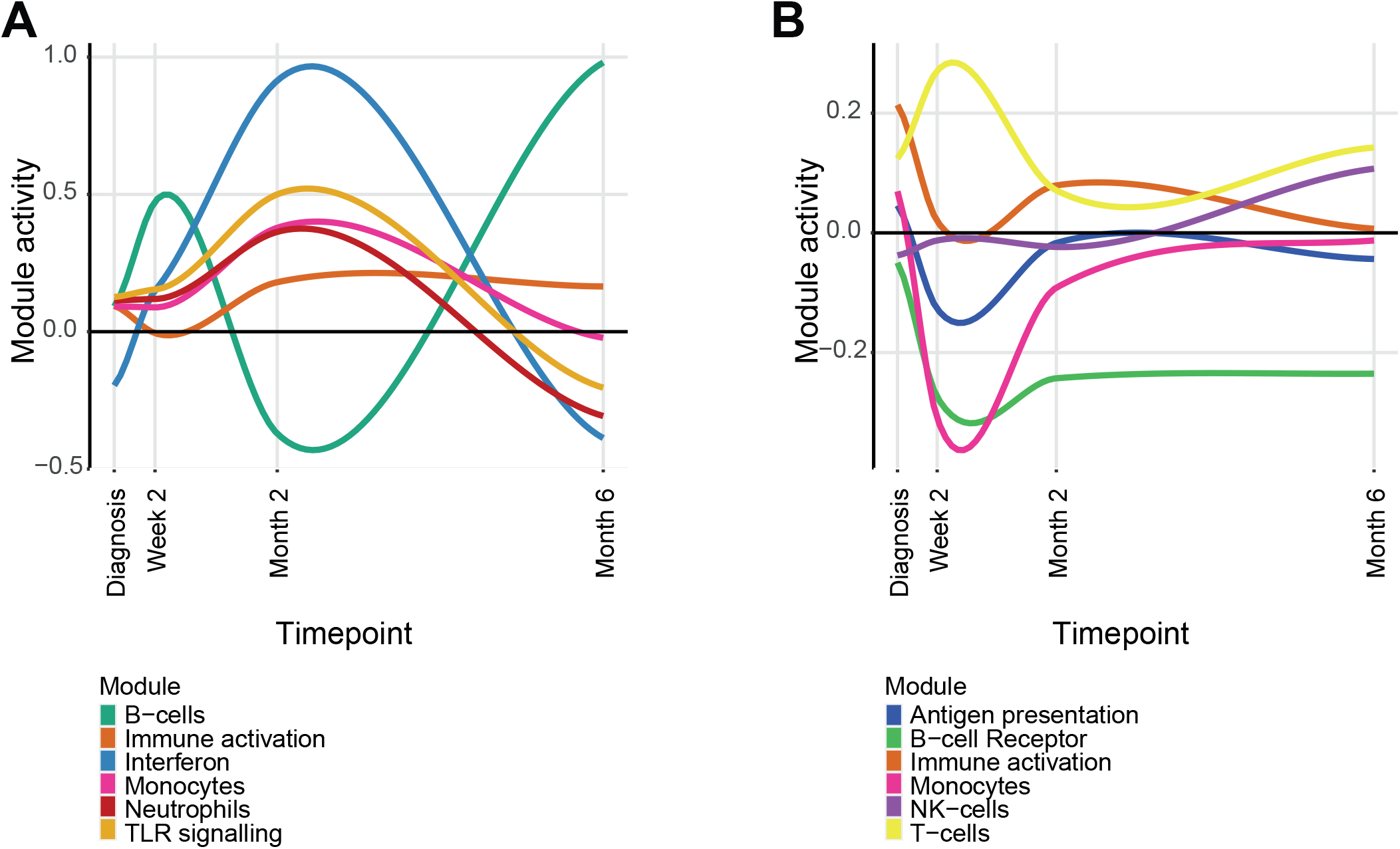
Modular activity of the most significant modules in TB-DM relative to TB-only in A) South Africa and B) Indonesia. Modular analysis was performed between TB-DM and TB-only patients and the most statistically significant were chosen (p-value <0.05). Modular activity calculated by summing the expression of genes within a module and dividing by the number of genes within that module.

### Impact of DM on TB treatment response using targeted gene expression profiling

We performed targeted profiling of TB-relevant immune gene expression in an expanded cohort from South Africa with more intensive sampling, using dcRT-MLPA(Supplementary Table S11A). At baseline, the overall MDP was similar in all study groups compared to healthy controls (Supplementary Figure S5A); Partial Least Squares – Discriminant Analysis separated DM-only patients from other groups including healthy controls, suggesting distinct genes are perturbed in DM-only (Supplementary Figure S5B; [25]). Gene expression was strongly correlated between TB-only and TB-DM, TB-preDM or TBrel-IH, but with some outlier genes which were affected by glycaemic status (Figure 5A). There were more DEGs relative to healthy controls in the TB-DM, TB-preDM and TBrel-IH groups than TB-only, at TB diagnosis and throughout treatment (Figure 5B). Normalisation of expression of genes such as *GNLY* and *GBP1* occurred by 2 weeks in the TB-only group but was delayed in TB-DM, TB-preDM and TBrel-IH. Results from targeted dcRT-MLPA analysis were thus in accordance with the global RNA-Seq analysis.

**Figure 5.**
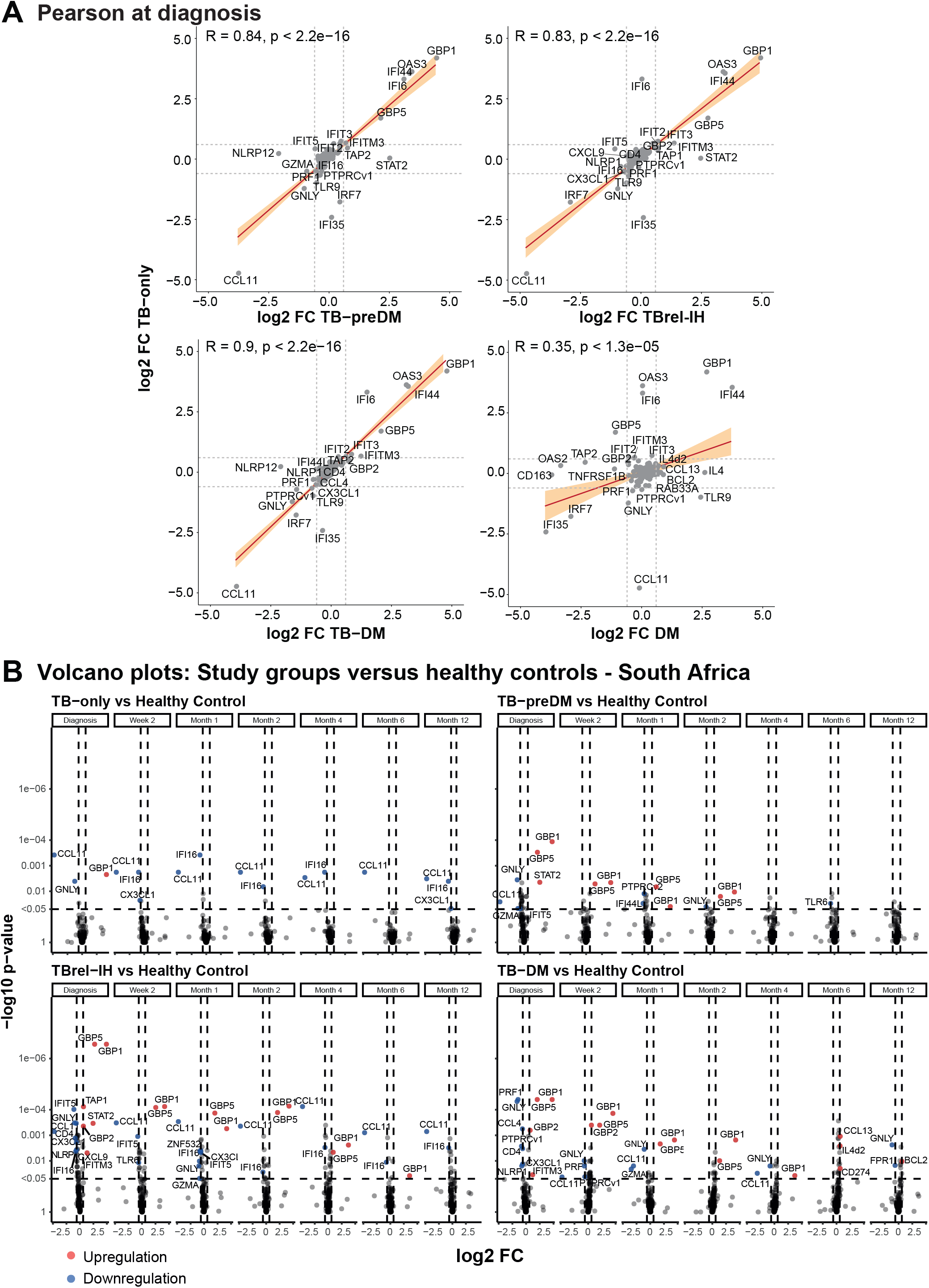
Gene expression profiles in TBrel-IH and TB-DM are not completely normalised to healthy control profiles at the end of TB treatment. **A)** Scatter plots representing Pearson correlations between expression of all genes in targeted dcRT-MLPA panel in TB patients relative to healthy controls (y-axes) versus the other study groups relative to healthy controls (x-axes), plotted as log_2_ FC. Red line corresponds to line of best fit and shaded bands indicate confidence intervals. Genes regulated log_2_ FC <-0.6 or > 0.6 are annotated. **B)** Differential Expression Analysis was performed on GAPDH-normalized log_2_-transformed targeted gene expression data of the South African cohort. Volcano plots representing DEGs at diagnosis and at different timepoints post TB treatment initiation of TB patients categorized based on their diabetes/glycaemia status compared to the healthy controls. The y-axis scales of all plots are harmonized per study group. P-values, -log_10_-transformed for better visualization, are plotted against log_2_ FC. Genes with p <0.05 and log_2_ FC <-0.6 or >0.6 were labelled as DEGs.

Longitudinal MDP analysis in the South African and Indonesian cohorts indicated the magnitude of transcriptomic response to TB treatment was dependent on diabetes/glycaemia with TB-DM patients displaying the largest gene expression perturbation over time (Figure 6A, Supplementary Figure S6A). Gene expression changes through treatment, identified by Linear Mixed Models, showed some consistency across TB groups, with the South African cohort exhibiting downregulation of *GBP5, GBP1*, and *IFITM3* (Figure 6B) and the Indonesian cohort showing downregulation of *GBP5* and *IFITM3* and upregulation of *GNLY* (Supplementary Figure 6B) from diagnosis to 6 months. Importantly, the number of upregulated DEGs in response to TB treatment increased with glycaemic index in both cohorts (South Africa: TB-only:6 DEGs, TB-preDM:10 DEGs, TBrel-IH:12 DEGs, TB-DM:14 DEGs; Indonesia: TB-only: 9 DEGs, TBrel-IH:13 DEGs, TB-DM:22 DEGs). Notably, no DEGs were detected between 6 and 12 months in the South African cohort, except for *GBP5* (p < 1e-10) in patients with TBrel-IH (Supplementary Figure S7).

**Figure 6.**
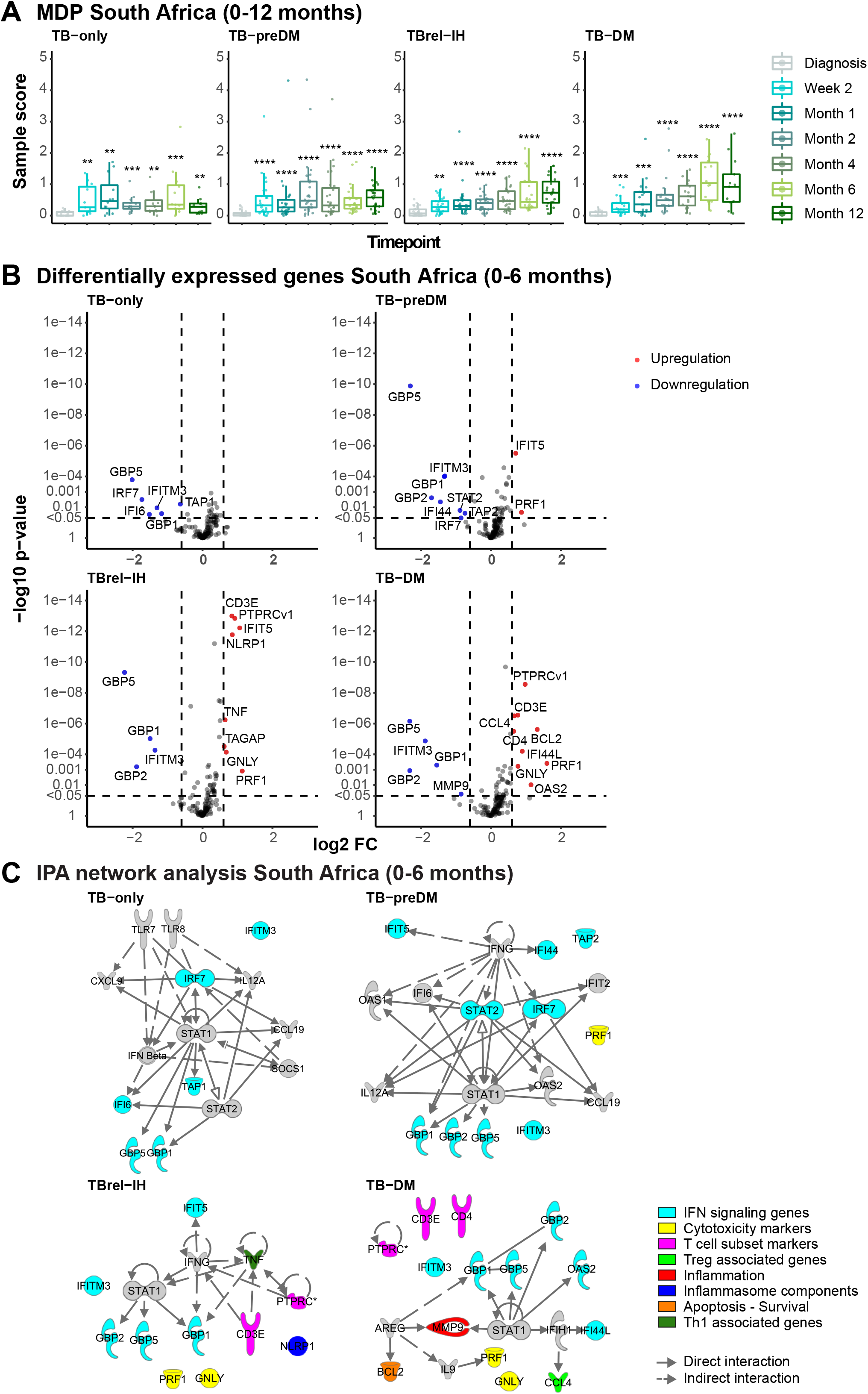
TB treatment response in TB patients is dependent on diabetes/glycaemia status. MDP and Differential Expression Analyses were performed on GAPDH-normalized log_2_-transformed targeted gene expression data of the South African cohort. (**A**) MDP analyses of the different study groups showing the impact of TB treatment on the overall gene perturbation over time. Samples of patients at diagnosis were used as baseline controls. (**B**) Volcano plots representing DEGs regulated during TB treatment of TB patients categorized based on their diabetes/glycaemia status. The y-axis scales of all plots are harmonized per study group. P-values, -log_10_-transformed for better visualization, are plotted against log_2_ FC. Genes with p <0.05 and log_2_ FC <-0.6 or >0.6 were labelled as DEGs. (**C**) IPA interactive network analyses of DEGs regulated during TB treatment. The various shapes of the nodes represent the functional classes of the gene products. Gene modules are indicated by distinctive colours.

Ingenuity Pathway Analysis showed the majority of treatment-response DEGs in TB-only and TB-preDM were Interferon-signaling genes (ISGs) (Figure 6C, Supplementary Figure S6C). In contrast, in TBrel-IH and TB-DM patients, although downregulation of ISGs through treatment was observed, the major change was upregulation of genes associated with adaptive immunity (T-cell subset markers, Th1-associated genes, Treg-associated genes, cytotoxicity markers). Overall, the dcRT-MLPA confirmed that although TB-associated gene profiles show similar pattern and rate of change in TB patients and TB-DM, the magnitude is different.

### Identification of a signature for TB treatment-response

As TB transcriptomic signatures were altered in people with DM or IH, we identified signatures with the highest classifying power to discriminate between patients at diagnosis and end of TB treatment irrespective of diabetes/glycaemia by pooling all TB patients, using logistic regression with lasso regularisation. Initially, signatures were developed in the South African and Indonesian cohorts separately (Table 2; Supplementary Table S12). The classifying capability of each signature against the training (AUC range: 0.73 – 1.0) and validation (AUC range: 0.69 – 0.92) cohorts for each clinical group was reasonably good (Figure 7A&B, Supplementary Figure 8A&B). To improve the classification performance and reduce cohort dependency, the datasets of both cohorts were pooled, and a combined two cohort 15-gene signature developed. This showed enhanced classification performance across the cohorts, with ROC analysis showing AUCs of 0.88 for TB-only, 0.96 for TBrel-IH and 0.85 for TB-DM, with excellent classification retained in individual cohorts (Figure 7C; Supplementary Figure 8C). The kinetic profiles of 6 representative genes are shown in Supplementary Figure S9.

**Table 2:** Gene expression signature predicting month 6 versus diagnosis, obtained by pooling the study groups and cohorts (South Africa + Indonesia)

| Gene | Module | Coefficient |
| --- | --- | --- |
| Intercept |  | -14.99967 |
| **BLR1** | G protein-coupled receptors | 0.07841 |
| **CCL13** | Chemokines | 0.40415 |
| **CCL4** | Treg associated genes | 0.66015 |
| **CD19** | Immune cell subset markers - B cells | 0.2371 |
| **CD3E** | T cell subset markers | 0.35071 |
| **CD4** | T cell subset markers | 0.00263 |
| **FCGR1A** | IFN signaling genes | -0.48224 |
| **FPR1** | Myeloid-associated genes | -0.1725 |
| **GBP5** | IFN signaling genes | -0.16957 |
| **IFIT5** | IFN signaling genes | 0.0172 |
| **NLRP1** | Inflammasome components | 0.3498 |
| **PTPRCv1** | T cell subset markers | 0.6625 |
| **TAP1** | IFN signaling genes | -0.51107 |
| **TNF** | Th1 associated genes | 0.06539 |
| **ZNF532** | Transcriptional regulators/activators | 0.06389 |

**Figure 7.**
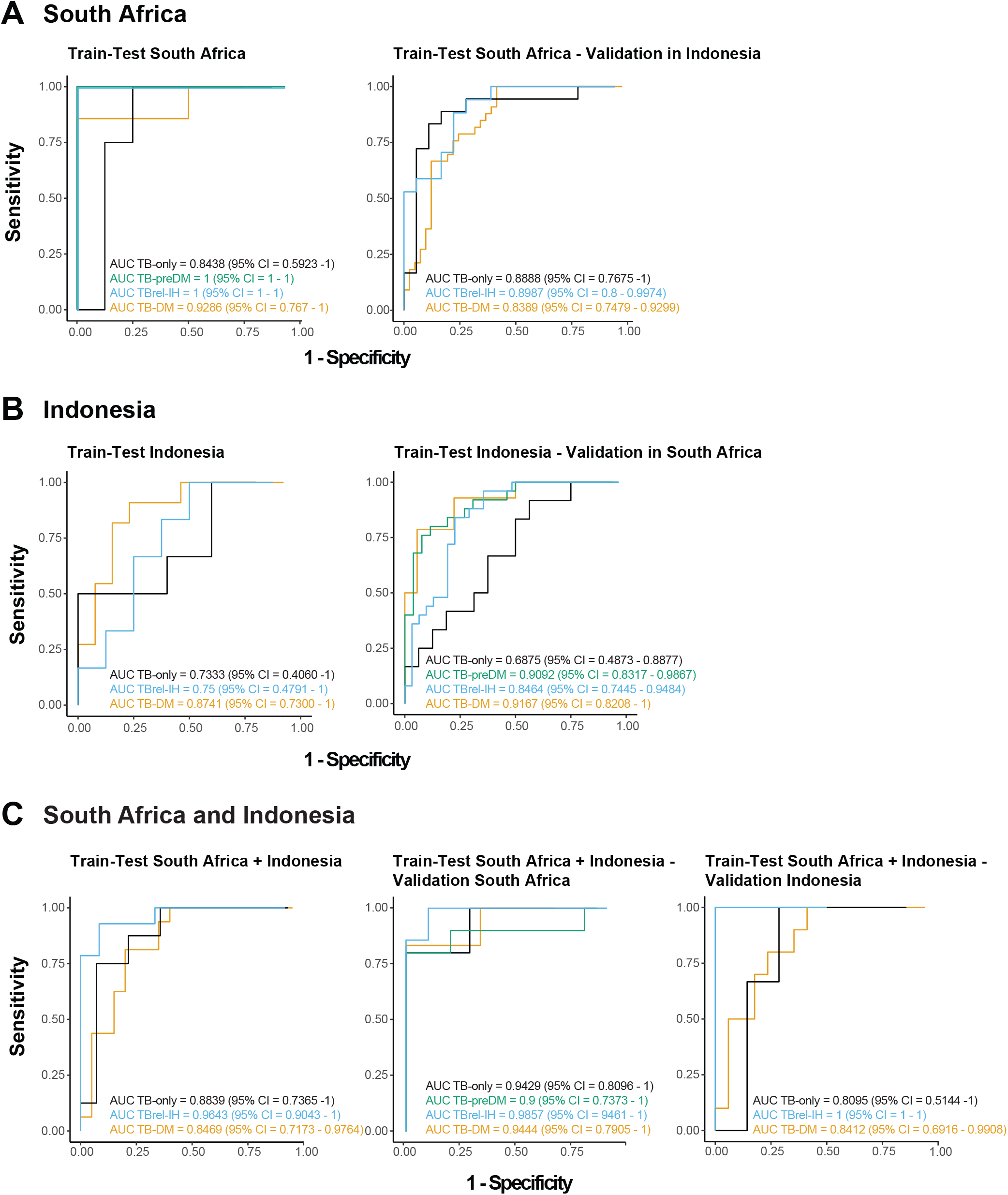
Identification of common host biomarker signatures associated with TB treatment response irrespective of population heterogeneity and diabetes/glycaemia severity. South African, Indonesian, or pooled cohort transcriptomic datasets of TB patients independent of their diabetes/glycaemia status were used to train the models. Receiver Operating Characteristic (ROC) curves (Sensitivity plotted against 1-Specificity) and Area Under the Curve (AUC) with 95% Confidence Intervals (CI) show the classifying performance of the trained models. (**A**) The model trained on 70% of the South African dataset was tested in the remaining 30% of the South African dataset split into the different TB study groups (left panel) and validated using the complete dataset of the Indonesian cohort split into the different TB study groups (right panel). (**B**) The model trained on 70% of the Indonesian dataset was tested in the remaining 30% of the Indonesian dataset split into the different TB study groups (left panel) and validated using the complete dataset of the South African cohort split into the different TB study groups (right panel). (**C**) The model trained on 70% of the pooled (South African and Indonesian) dataset was tested in the remaining 30% of the pooled dataset split into the different TB study groups that both cohorts have in common (left panel) and validated using the complete dataset of the South African cohort split into the different TB study groups (middle panel) or the complete dataset of the Indonesian cohort split into the different TB study groups (right panel).

## Discussion

In this longitudinal analysis of blood transcriptomes, excessive gene expression perturbation previously described at TB diagnosis [13, 25] continued throughout six months of TB treatment in pulmonary TB patients with diabetes co-morbidity. However, qualitatively and kinetically similar changes occurred in patients with or without diabetes, suggesting prolonged TB treatment might be sufficient to restore normal transcriptomes. TB patients with either pre-diabetes or TB-related IH also exhibited greater magnitudes of gene expression perturbation throughout treatment, similar to patients with diagnosed diabetes. The overall consistency in change of gene expression through treatment, irrespective of glycaemic index, enabled derivation of accurate predictive models of TB treatment response, which could be used effectively in populations with or without diabetes.

Diabetes has a negative effect on TB treatment outcomes [5, 6], for unclear reasons. One explanation could be a qualitatively different immune response in diabetes, leaving people persistently susceptible to bacterial replication and disease reactivation. An alternative explanation is that excessive inflammation and immune activation at diagnosis in TB-DM means patients require longer or, more likely, different treatment to reach the same endpoint as people with uncomplicated TB, so that they are not left susceptible to TB recurrence. Our data support the latter model, as all gene clusters differentially expressed between clinical groups exhibited similar changes, but of different magnitude. Bronchial spread often persists beyond treatment initiation, with new or expanding cavities appearing on PET-CT scans 4 weeks into treatment in one-fifth of pulmonary TB patients [32].

Plausibly, increased ongoing bacterial spread in patients with diabetes co-morbidity causes persistent pro-inflammatory responses: the peripheral transcriptome correlates with lung inflammatory activity in TB patients [33]. Restoration of normal transcriptomes, and presumably improved lung resolution, could potentially also be achieved by co-administration of host-directed therapy alongside standard treatment. Therapy which dampens pro-inflammatory responses, such as corticosteroids or matrix metalloproteinase inhibitors [34], would have added benefit by reducing lung damage, which often persists after microbiological cure [35]. Anti-hyperglycaemic therapy, such as metformin, leads to more balanced, less inflammatory responses to *M*.*tuberculosis* [36], and has been suggested as adjunctive therapy for TB, particularly in patients with diabetes [37]. Our transcriptomic data suggest that patients with either pre-diabetes or TB-related IH would also benefit from prolonged or adjunctive host-directed therapy, in alignment with observed worse TB treatment outcomes in people with transient hyperglycaemia [38].

The ability to monitor TB treatment and predict outcome would be beneficial for clinical management. We show that transcriptomic models can be derived from host blood which reflect TB treatment-response irrespective of glycaemia. The best models include genes involved in interferon signalling, known to be suppressed at TB diagnosis in TB-DM patients [25], which we found were enhanced mid-way through treatment but did eventually resolve by 6 months. People protected against TB development display balanced prostaglandin 2 and lipoxin expression in lungs, preventing TB disease progression following infection [39]. Drugs which target 5-lipoxygenase restrict lung pathology and reduce bacterial replication in murine models, by lowering the type 1 interferon response; the increases through treatment in the TB-DM cohort may relate to sustained infection and accompanying inflammation. In TB-DM patients the inflammation-related genes resolved more linearly through TB treatment, but remained elevated to the end of TB treatment, persisting until 12 months post-diagnosis in the South African cohort. In future studies, it would be important to test whether prolonged treatment with standard therapy impacts blood transcriptomes beyond the 6 month time point. Increased doses of anti-TB drugs might also lead to better treatment outcomes in TB-DM. In a companion paper (van Doorn *et al*, [40] unpublished), transcriptomic signatures indicative of treatment outcome have been derived that can be used in patients with either DM or IH. Together, these papers show that signatures related to poor TB outcome are distinct from the excessive and prolonged inflammation observed in TB-DM.

These findings further illustrate how comorbidity with diabetes affects the host response to *M*.*tuberculosis* infection, and how a better understanding of these interactions could be exploited to reduce poor TB treatment outcomes associated with TB and diabetes comorbidity.

## Supporting information

TANDEM_Full_Consortium

## Data Availability

RNASeq data will be available online at NCBI GEO once manuscript is published, Accession number GSE193978. dcRT-MLPA data are available upon reasonable request to the authors.

## Acknowledgments

We are grateful to all the study participants for blood and data donations. We acknowledge□Bahram Sanjabi, Desiree Brandenburg-Weening, and Pieter van der□Vlies□for assistance with the RNA-Seq,□Evelien□Temminck□for□providing technical assistance with□dcRT-MLPA□experiments, and J□Erni□Durdevic□for providing statistical□and machine learning□advice.□

## References

1. Koesoemadinata RC, McAllister SM, Soetedjo NNM, et al. Latent TB infection and pulmonary TB disease among patients with diabetes mellitus in Bandung, Indonesia. Trans R Soc Trop Med Hyg 2017; 111(2): 81–9.

2. Al-Rifai RH, Pearson F, Critchley JA, Abu-Raddad LJ. Association between diabetes mellitus and active tuberculosis: A systematic review and meta-analysis. PLoS One 2017; 12(11): e0187967.

3. Jeon CY, Murray MB. Diabetes mellitus increases the risk of active tuberculosis: a systematic review of 13 observational studies. PLoS Med 2008; 5(7): e152.

4. Noubiap JJ, Nansseu JR, Nyaga UF, et al. Global prevalence of diabetes in active tuberculosis: a systematic review and meta-analysis of data from 2.3 million patients with tuberculosis. Lancet Glob Health 2019; 7(4): e448–e60.

5. Baker MA, Harries AD, Jeon CY, et al. The impact of diabetes on tuberculosis treatment outcomes: a systematic review. BMC Med 2011; 9: 81.

6. Huangfu P, Ugarte-Gil C, Golub J, Pearson F, Critchley J. The effects of diabetes on tuberculosis treatment outcomes: an updated systematic review and meta-analysis. Int J Tuberc Lung Dis 2019; 23(7): 783–96.

7. International Diabetes Federation. IDF Diabetes Atlas Ninth Edition, 2019 2017.

8. American Diabetes Association. Diagnosis and classification of diabetes mellitus. Diabetes Care 2010; 33 Suppl 1: S62–9.

9. Dungan KM, Braithwaite SS, Preiser JC. Stress hyperglycaemia. Lancet 2009; 373(9677): 1798–807.

10. Boillat-Blanco N, Ramaiya KL, Mganga M, et al. Transient Hyperglycemia in Patients With Tuberculosis in Tanzania: Implications for Diabetes Screening Algorithms. J Infect Dis 2016; 213(7): 1163–72.

11. Yoon YS, Jung JW, Jeon EJ, et al. The effect of diabetes control status on treatment response in pulmonary tuberculosis: a prospective study. Thorax 2017; 72(3): 263–70.

12. Ronacher K, Joosten SA, van Crevel R, Dockrell HM, Walzl G, Ottenhoff TH. Acquired immunodeficiencies and tuberculosis: focus on HIV/AIDS and diabetes mellitus. Immunol Rev 2015; 264(1): 121–37.

13. Prada-Medina CA, Fukutani KF, Pavan Kumar N, et al. Systems Immunology of Diabetes-Tuberculosis Comorbidity Reveals Signatures of Disease Complications. Sci Rep 2017; 7(1): 1999.

14. Ronacher K, Chegou NN, Kleynhans L, et al. Distinct serum biosignatures are associated with different tuberculosis treatment outcomes. Tuberculosis (Edinb) 2019; 118: 101859.

15. Kumar NP, Banurekha VV, Nair D, et al. Coincident pre-diabetes is associated with dysregulated cytokine responses in pulmonary tuberculosis. PLoS One 2014; 9(11): e112108.

16. Kumar NP, Fukutani KF, Shruthi BS, et al. Persistent inflammation during anti-tuberculosis treatment with diabetes comorbidity. Elife 2019; 8.

17. Kumar NP, Moideen K, Sivakumar S, et al. Modulation of dendritic cell and monocyte subsets in tuberculosis-diabetes co-morbidity upon standard tuberculosis treatment. Tuberculosis (Edinb) 2016; 101: 191–200.

18. Kumar NP, Moideen K, Viswanathan V, Kornfeld H, Babu S. Effect of standard tuberculosis treatment on naive, memory and regulatory T-cell homeostasis in tuberculosis-diabetes co-morbidity. Immunology 2016; 149(1): 87–97.

19. Berry MP, Graham CM, McNab FW, et al. An interferon-inducible neutrophil-driven blood transcriptional signature in human tuberculosis. Nature 2010; 466(7309): 973–7.

20. Cliff JM, Lee JS, Constantinou N, et al. Distinct phases of blood gene expression pattern through tuberculosis treatment reflect modulation of the humoral immune response. J Infect Dis 2013; 207(1): 18–29.

21. Kaforou M, Wright VJ, Oni T, et al. Detection of tuberculosis in HIV-infected and -uninfected African adults using whole blood RNA expression signatures: a case-control study. PLoS Med 2013; 10(10): e1001538.

22. Ottenhoff TH, Dass RH, Yang N, et al. Genome-wide expression profiling identifies type 1 interferon response pathways in active tuberculosis. PLoS One 2012; 7(9): e45839.

23. Bloom CI, Graham CM, Berry MP, et al. Detectable changes in the blood transcriptome are present after two weeks of antituberculosis therapy. PLoS One 2012; 7(10): e46191.

24. Thompson EG, D. Y, Malherbe ST, et al. Host blood RNA signatures predict the outcome of tuberculosis treatment. Tuberculosis 2017; 107: 48–58.

25. Eckold C, Kumar V, Weiner J, et al. Impact of Intermediate Hyperglycemia and Diabetes on Immune Dysfunction in Tuberculosis. Clin Infect Dis 2021; 72(1): 69–78.

26. Ugarte-Gil C, Alisjahbana B, Ronacher K, et al. Diabetes Mellitus Among Pulmonary Tuberculosis Patients From 4 Tuberculosis-endemic Countries: The TANDEM Study. Clin Infect Dis 2020; 70(5): 780–8.

27. Ruslami R, Koesoemadinata RC, Soetedjo NNM, et al. The effect of a structured clinical algorithm on glycemic control in patients with combined tuberculosis and diabetes in Indonesia: A randomized trial. Diabetes research and clinical practice 2021; 173: 108701.

28. Grint D, Alisjhabana B, Ugarte-Gil C, et al. Accuracy of diabetes screening methods used for people with tuberculosis, Indonesia, Peru, Romania, South Africa. Bull World Health Organ 2018; 96(11): 738–49.

29. Nueda MJ, Tarazona S, Conesa A. Next maSigPro: updating maSigPro bioconductor package for RNA-seq time series. Bioinformatics 2014; 30(18): 2598–602.

30. Joosten SA, Goeman JJ, Sutherland JS, et al. Identification of biomarkers for tuberculosis disease using a novel dual-color RT-MLPA assay. Genes Immun 2012; 13(1): 71–82.

31. Dennis G, Jr., Sherman BT, Hosack DA, et al. DAVID: Database for Annotation, Visualization, and Integrated Discovery. Genome Biol 2003; 4(5): P3.

32. Chen RY, Yu X, Smith B, et al. Radiological and functional evidence of the bronchial spread of tuberculosis: an observational analysis. The Lancet Microbe 2021; 2(10): e518–e26.

33. Odia T, Malherbe ST, Meier S, et al. The Peripheral Blood Transcriptome Is Correlated With PET Measures of Lung Inflammation During Successful Tuberculosis Treatment. Frontiers in immunology 2020; 11: 596173.

34. Young C, Walzl G, Du Plessis N. Therapeutic host-directed strategies to improve outcome in tuberculosis. Mucosal Immunology 2020; 13(2): 190–204.

35. Krug S, Parveen S, Bishai WR. Host-Directed Therapies: Modulating Inflammation to Treat Tuberculosis. Frontiers in immunology 2021; 12: 660916.

36. Lachmandas E, Eckold C, Bohme J, et al. Metformin Alters Human Host Responses to Mycobacterium tuberculosis in Healthy Subjects. J Infect Dis 2019; 220(1): 139–50.

37. Singhal A, Jie L, Kumar P, et al. Metformin as adjunct antituberculosis therapy. Science translational medicine 2014; 6(263): 263ra159.

38. Liu Q, You N, Pan H, et al. Glycemic Trajectories and Treatment Outcomes of Patients with Newly Diagnosed Tuberculosis: A Prospective Study in Eastern China. Am J Respir Crit Care Med 2021; 204(3): 347–56.

39. Mayer-Barber KD, Andrade BB, Oland SD, et al. Host-directed therapy of tuberculosis based on interleukin-1 and type I interferon crosstalk. Nature 2014; 511(7507): 99–103.

40. van Doorn CLR, Eckold C, Ronacher K, et al. Transcriptional profiles predict treatment outcome in patients with tuberculosis and diabetes at diagnosis and at two weeks after initiation of anti-tuberculosis treatment. MedRxiv 2022; (MEDRXIV/2022/269796).

