## Supplementary Methods for "Impaired resolution of blood transcriptomes through tuberculosis treatment with diabetes comorbidity"

*RNASeq data analysis*

RNA Samples processed for RNA-Seq analysis were quality-assessed by LabChip GX HiSens RNA system (PerkinElmer). Total RNA samples were processed using the poly-A tail Bioscientific NEXTflex-Rapid-Directional mRNA-seq method with the Caliper SciClone to generate libraries, which were single-end sequenced using the NextSeq500 High Output kit V2 (Illumina) for 75 cycles. Data are deposited in the NCBI-GEO database, accession number GSE193978. STAR (v2.5.1b) [1] was used to align the sequence data from FASTQ files, to the Human g1kv37 reference genome, and quality control performed with FastQC [2]. Downstream analysis was performed in R [3]. HTseq-count (v0.61) was used for transcript quantification [4]. Lowly expressed transcripts for which counts did not exceed 50 across all samples were removed. Data were normalised using R package DESeq2 (1.30.0) [5]. For the MaSigPro [6] analysis, due to the number of timepoints, a quadratic regression model (degrees of freedom = 2) was executed. False discovery correction was done using Benjamini-Hochberg method with an adjusted p-value of < 0.05 deemed significant. Molecular Degree of Perturbation (MDP) analysis was performed using the R package mdp [7]. The R package tmod [8] was used to perform modular analysis on the genes found to be differentially expressed between clinical groups using its HGtest function against all genes as a background. Modules with an adjusted p-value <0.05 were deemed significant. Modular activity was calculated by summing the differential expression of genes in TB/DM relative to TB-only within a module and then dividing by the number of genes within that module. The g:profiler webtool [9] was used for gene ontology and pathway analyses gene lists.

*Reverse-Transcriptase Multiplex Ligation-dependent Probe Amplification (dcRT-MLPA)*

Reverse-Transcriptase Multiplex Ligation-dependent Probe Amplification (dcRT-MLPA) was performed using the SALSA MLPA kit (MRC-Holland) as described elsewhere [10]. RT primers and half-probes were designed by Leiden University Medical Centre (LUMC, Leiden, the Netherlands) and comprised sequences for 4 housekeeping genes and 144 selected key immune-related genes to profile specific compartments of the human immune response (Supplementary Table S1). Briefly, 125 ng RNA was reverse transcribed to cDNA by incubation at 37°C for 15 min using Moloney Murine Leukemia Virus (M-MLV) reverse transcriptase (Promega, Leiden, The Netherlands), gene-specific RT-primers (Sigma-Aldrich, Zwijndrecht, The Netherlands), followed by inactivation of the enzyme by heating at 98⁰C for 2 min. The left- and right-hand half probes were hybridized to the cDNA at 60⁰C overnight, followed by ligation at 54⁰C for 15 minutes using ligase-65 (MRC-Holland), and inactivation by heating at 98⁰C for 5 min. Ligated probes were amplified by PCR (33 cycles at 95⁰C for 30 seconds, 58⁰C for 30 seconds and 72⁰C for 60 seconds, followed by one cycle at 72⁰C for 20 minutes). PCR products were 1:10 diluted in Highly deionized (Hi-Di) formamide (ThermoFisher) containing 400HD Rhodamine X (ROX) fluorophore size standard (ThermoFisher). PCR products were denatured at 95⁰C for 5 min, stored immediately at 4⁰C and analyzed on an Applied Biosystems 3730 capillary sequencer in GeneScan mode (BaseClear, Leiden, The Netherlands). Trace data were analyzed using GeneMapper software 5 (Applied Biosystems, Warrington, UK). The areas of each assigned peak (arbitrary units) were exported for analysis in R (version 3.6.3). Data were normalized to housekeeping gene glyceraldehyde 3-phosphate dehydrogenase (GAPDH) and signals below the threshold value for noise cutoff in GeneMapper (log2 transformed peak area 7.64) were assigned the threshold value for noise cutoff.

dcRT-MLPA data were analysed to identify differentially expressed genes (DEGs) between groups at diagnosis by the non-parametric Mann-Whitney U-test with Benjamini-Hochberg correction for multiple testing. Ingenuity Pathway Analysis (IPA-60467501) (QIAGEN, Hilden, Germany) was used to explore interactive networks between the DEGs. Molecular Degree of Perturbation (MDP) analysis (mdp R package) [7], Partial Least Squares – Discriminant Analysis (PLS-DA) (mixOmics R package) [11] and Pearson correlations of gene expression (log2 FC) versus healthy control were performed in R version 4.0.2. Longitudinal changes in gene expression levels from diagnosis (baseline) to 6 months (Indonesian cohort) or 12 months (South African cohort) were assessed by means of linear mixed models for repeated measurements over time. Models were fitted to base 2 logarithm transformed measurements using lme4 package in R using the lmer function [12]. Group-time interactions were included as fixed effects and the patients ID-time interactions were included as random effects. For the South African cohort, we forced a b-spline at 6 months, which enabled us to identify altered gene expressions during treatment (0-6 months) as well as altered gene expression after treatment (6-12 months). Time was coded as 0 for the first timepoint (diagnosis) and as a continuous variable for the time difference between the two time points. P-values were adjusted for multiple testing using the False Discovery Rate (FDR) method of Benjamini-Hochberg [13]. An adjusted p-value < 0.05 and a log2-fold change (FC) < -0.6 and > 0.6 were set as threshold for the identification of differentially expressed genes (DEGs). Genes that were below the detection limit in >90% of the samples per cohort were excluded from analysis. Signatures with the best discriminatory capability were identified using logistic regression with lasso regularization (glmnet R package) [14]. Leave-One-Out Cross Validation (LOOCV) and Train-Test Split (TTS) were used to assess the performance of the trained regression models. The classifying performance of the models were assessed by evaluating sensitivity, specificity, Receiver Operating Characteristic (ROC) curve, and Area Under the ROC Curve (AUC) with 95% Confidence Interval (CI), and box-and-whiskers-plots representing the predicted probability for each class were used to evaluate the classifying performance of the models.

**Supplementary References**

1.	Dobin A, Davis CA, Schlesinger F, et al. STAR: ultrafast universal RNA-seq aligner. Bioinformatics **2013**; 29(1): 15-21.

2.	Andrews S. FastQC: a quality control tool for high throughput sequence data. Available at: <http://www.bioinformatics.babraham.ac.uk/projects/fastqc>.

3.	R Core Team. R: A language and environment for statistical computing. Vienna, Austria: R Foundation for Statistical Computing, **2013**.

4.	Anders S, Pyl PT, Huber W. HTSeq--a Python framework to work with high-throughput sequencing data. Bioinformatics **2015**; 31(2): 166-9.

5.	Love MI, Huber W, Anders S. Moderated estimation of fold change and dispersion for RNA-seq data with DESeq2. Genome Biol **2014**; 15(12): 550.

6.	Nueda MJ, Tarazona S, Conesa A. Next maSigPro: updating maSigPro bioconductor package for RNA-seq time series. Bioinformatics **2014**; 30(18): 2598-602.

7.	Lever M RP, Nakaya H. mdp: Molecular Degree of Perturbation calculates scores for transcriptome data samples based on their perturbation from controls. R package version 1.12.0. Available at: <https://mdp.sysbio.tools/>.

8.	Weiner 3rd J, Domaszewska T. tmod: an R package for general and multivariate enrichment analysis. PeerJ Preprints **2016**; 4: e2420v1

9.	Raudvere U, Kolberg L, Kuzmin I, et al. g:Profiler: a web server for functional enrichment analysis and conversions of gene lists (2019 update). Nucleic Acids Res **2019**; 47(W1): W191-W8.

10.	Joosten SA, Goeman JJ, Sutherland JS, et al. Identification of biomarkers for tuberculosis disease using a novel dual-color RT-MLPA assay. Genes Immun **2012**; 13(1): 71-82.

11.	Kim-Anh Le Cao FR, Ignacio Gonzalez, Sebastien Dejean. mixOmics: Omics Data Integration Project. R package version 6.1.1. Available at: <https://CRAN.R-project.org/package=mixOmics>.

12.	Bates D, Mächler M, Bolker BM, Walker SC. Fitting linear mixed-effects models using lme4. Journal of Statistical Software **2015**; 67.

13.	Benjamini Y, Hochberg Y. Controlling the false discovery rate: a practical and powerful approach to multiple testing. Journal of the Royal statistical society: series B (Methodological) **1995**; 57(1): 289-300.

14.	Friedman J HT, Tibshirani R. Regularization Paths for Generalized Linear Models via Coordinate Descent. Journal of Statistical Software **2010**; 33(1), 1–22.
