## Supplementary Figures for "Impaired resolution of blood transcriptomes through tuberculosis treatment with diabetes comorbidity"

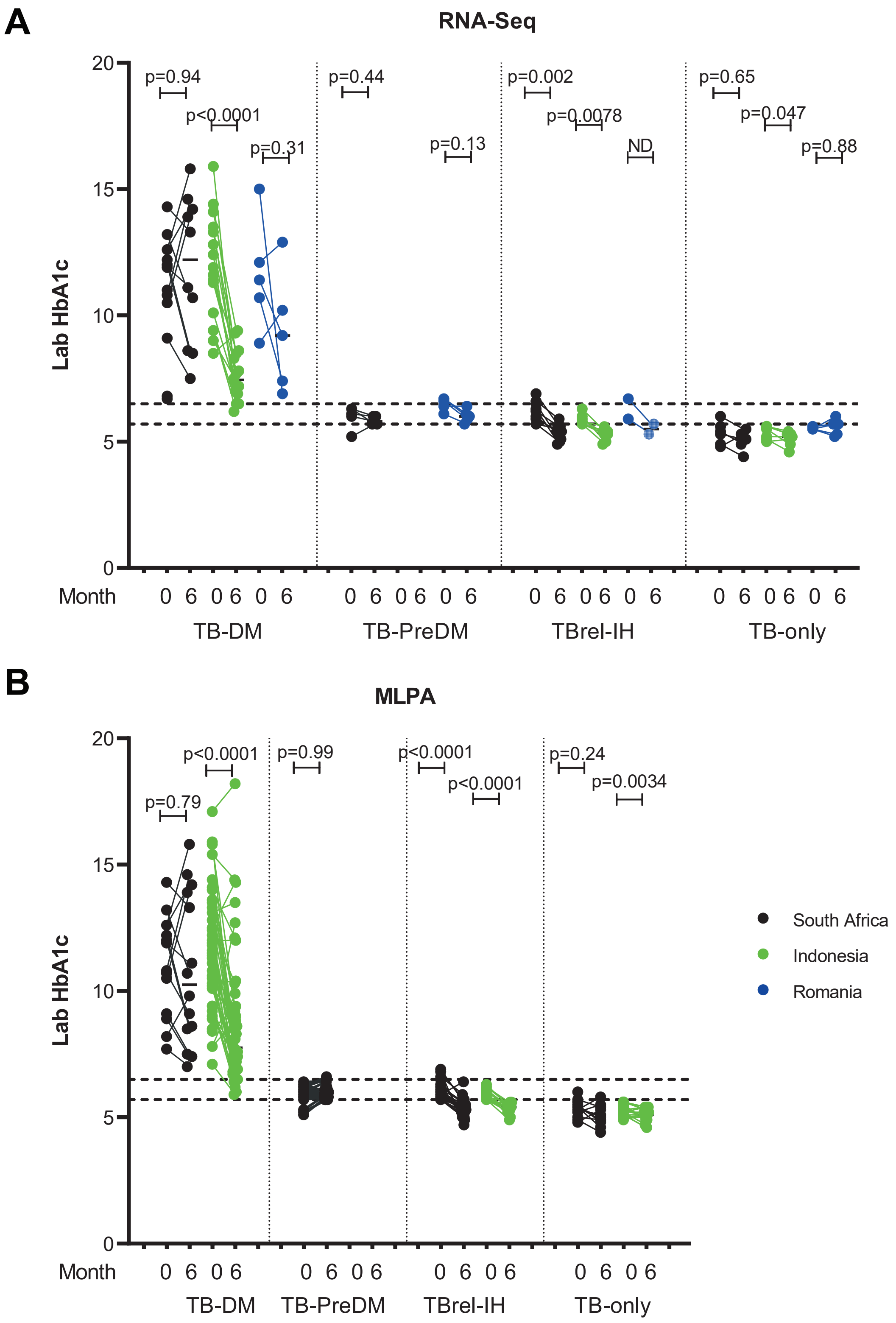


**Supplementary Figure 1. Changes in HbA1c in pulmonary TB patients through TB drug treatment in people with concomitant diabetes, pre-diabetes or TB-related intermediate hyperglycemia.** Laboratory HbA1c recorded at month 0, prior to initiation of treatment, and at the end of treatment at month 6 in patients included in A) RNA-Seq analysis and B) IMLPA analysis, for cohorts from South Africa (Black), Indonesia (Green) and Romania (Blue). The dotted lines show the threshold for diagnosis of diabetes (6.5%) and pre-diabetes (5.7%) [8]. Timepoints in each cohort were compared by Wilcoxon signed rank test.

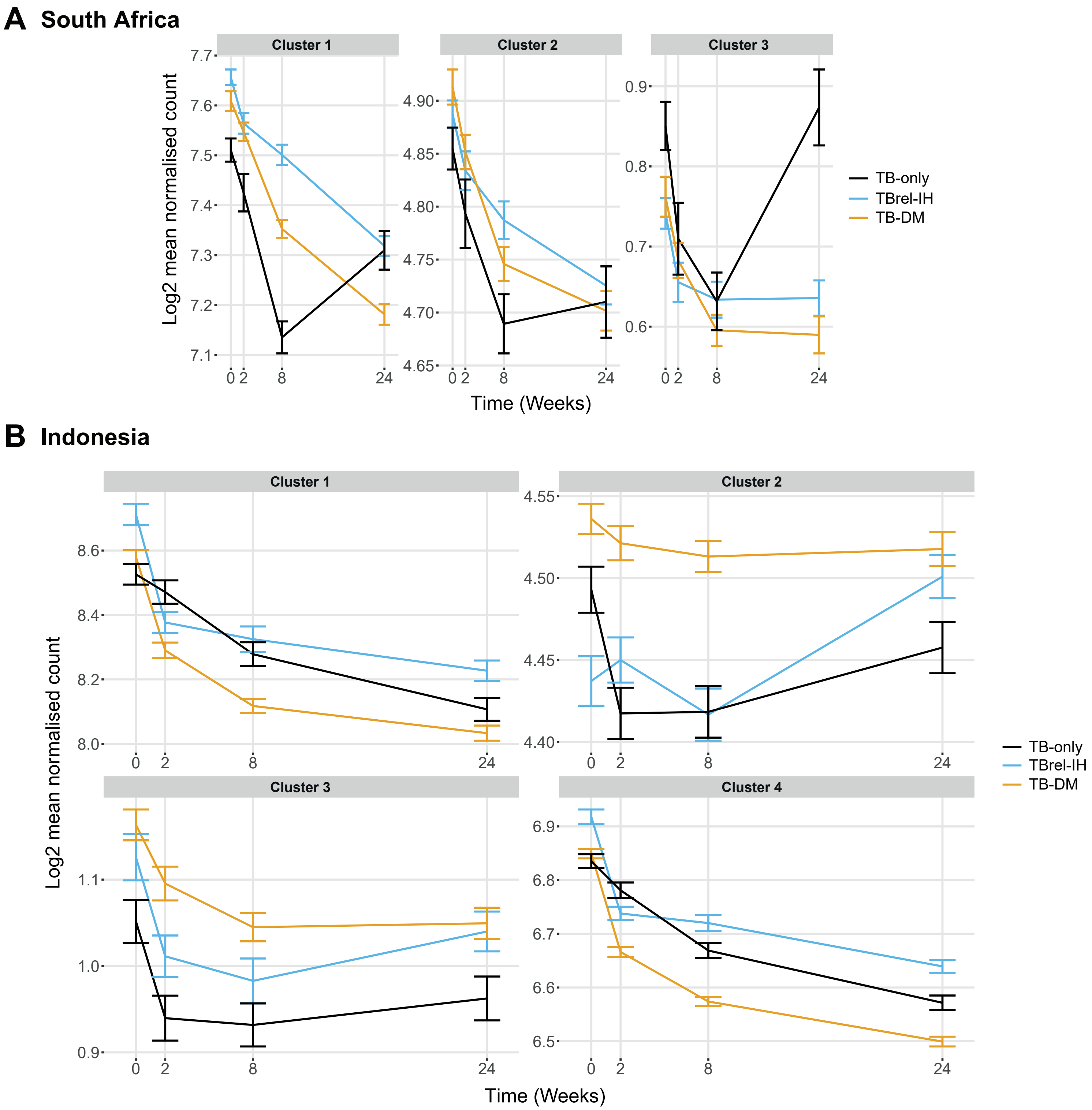


**Supplementary Figure 2. MaSigPro analysis of change in gene expression through TB treatment in blood samples from patients in A) South Africa and B) Indonesia.** MaSigPro identified genes that behaved similarly through time and between patient groups using hierarchical clustering. Results are shown for log-transformed normalised transcript counts for the TB-only, TB-DM and the combined TB-PreDM and TBrel-IH groups. Bars show mean ± 1 SEM. Data were filtered to remove lowly abundant transcripts prior to analysis. NB: There were insufficient study participants to conduct an analysis separately for the Romanian population.

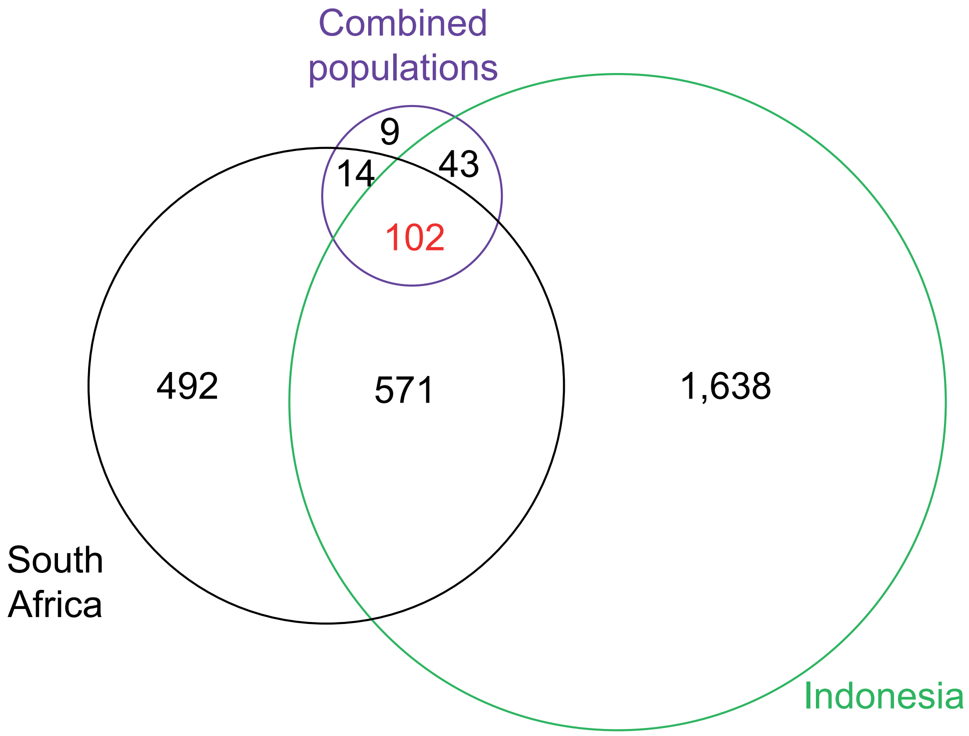


**Supplementary Figure 3. Comparison of results from MaSigPro analyses for individual and combined populations.** Genes which were identified as behaving differently between TB-DM, TBrel-IH/TB-PreDM or TB-only in South Africa (Black), Indonesia (Green) or combined populations from Romania, South Africa and Indonesia (Purple) in MaSigPro analyses (Figures 2, S2; Supplementary Tables S4, S5, S6) were compared. The Venn Diagram shows the number of identified genes which overlap between analyses.

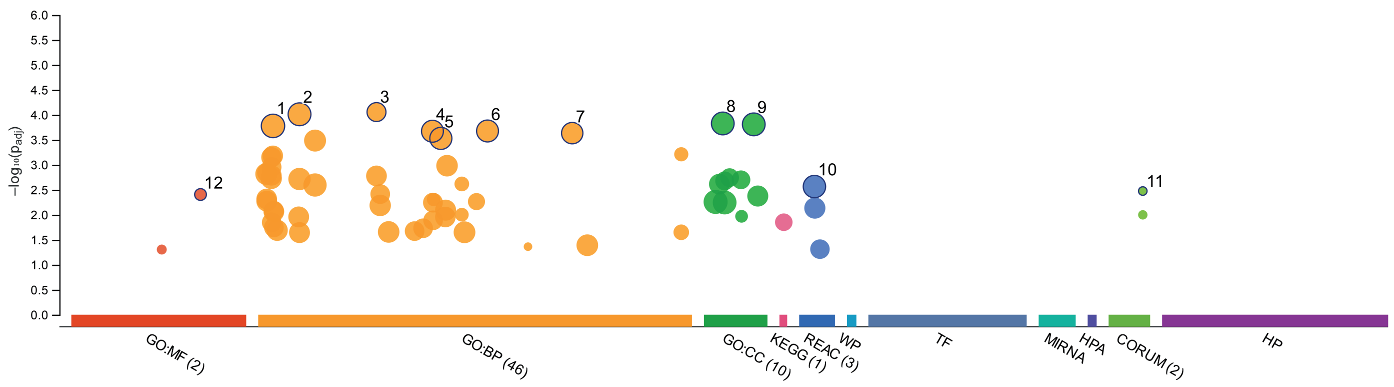


| ID | Source | Term ... | Term Name | $p_{adj}$ (query_1) |
|---|---|---|---|---|
| 1 | GO:BP | GO:0002376 | immune system process | $1.664 \times 10^{-4}$ |
| 2 | GO:BP | GO:0006955 | immune response | $9.729 \times 10^{-5}$ |
| 3 | GO:BP | GO:0031349 | positive regulation of defense response | $8.788 \times 10^{-5}$ |
| 4 | GO:BP | GO:0043207 | response to external biotic stimulus | $2.126 \times 10^{-4}$ |
| 5 | GO:BP | GO:0044419 | biological process involved in interspecies interacti... | $2.954 \times 10^{-4}$ |
| 6 | GO:BP | GO:0051707 | response to other organism | $2.101 \times 10^{-4}$ |
| 7 | GO:BP | GO:0098542 | defense response to other organism | $2.320 \times 10^{-4}$ |
| 8 | GO:CC | GO:0031410 | cytoplasmic vesicle | $1.486 \times 10^{-4}$ |
| 9 | GO:CC | GO:0097708 | intracellular vesicle | $1.548 \times 10^{-4}$ |
| 10 | REAC | REAC:R-HSA-16... | Immune System | $2.721 \times 10^{-3}$ |
| 11 | CORUM | CORUM:6826 | Calprotectin heterotetramer | $3.346 \times 10^{-3}$ |
| 12 | GO:MF | GO:0050786 | RAGE receptor binding | $3.919 \times 10^{-3}$ |

| | |
|---|---|
| **version** | e104_eg51_p15_3922dba |
| **date** | 13/12/2021, 13:09:13 |
| **organism** | hsapiens |

g:Profiler

**Supplementary Figure 4. G:profiler analysis of 102 gene MSP core gene list, identifying top Gene Ontology (GO) molecular function (MF), biological process (BP) and cellular component (CC) as well as biological pathways in KEGG and Reactome databases.**

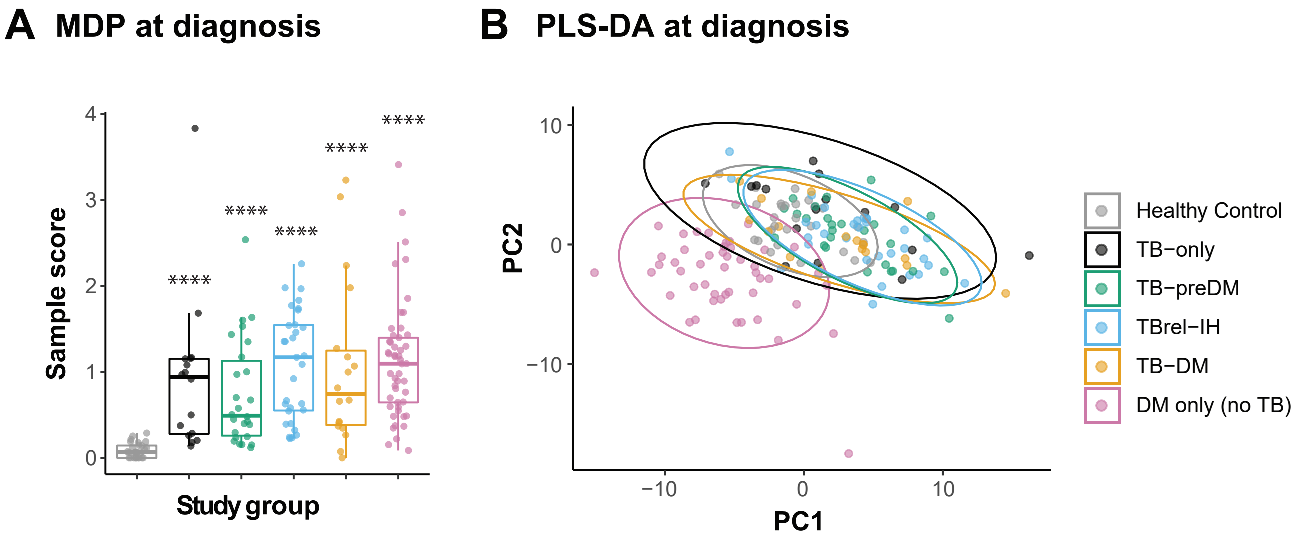


**Supplementary Figure S5. Gene expression profiles in TB patients is dominant over the gene perturbation caused by DM.** Molecular Degree of Perturbation (MDP) and Partial Least Squares – Discriminant Analysis (PLS-DA) were performed on GAPDH-normalized log2-transformed targeted gene expression data of the South African cohort. (A) MDP analysis of the different study groups showing the impact of TB and DM on the overall gene perturbation at diagnosis. Samples of healthy individuals were used as controls. (B) PLS-DA analysis of the different study groups at diagnosis. Samples of healthy individuals were used as controls.

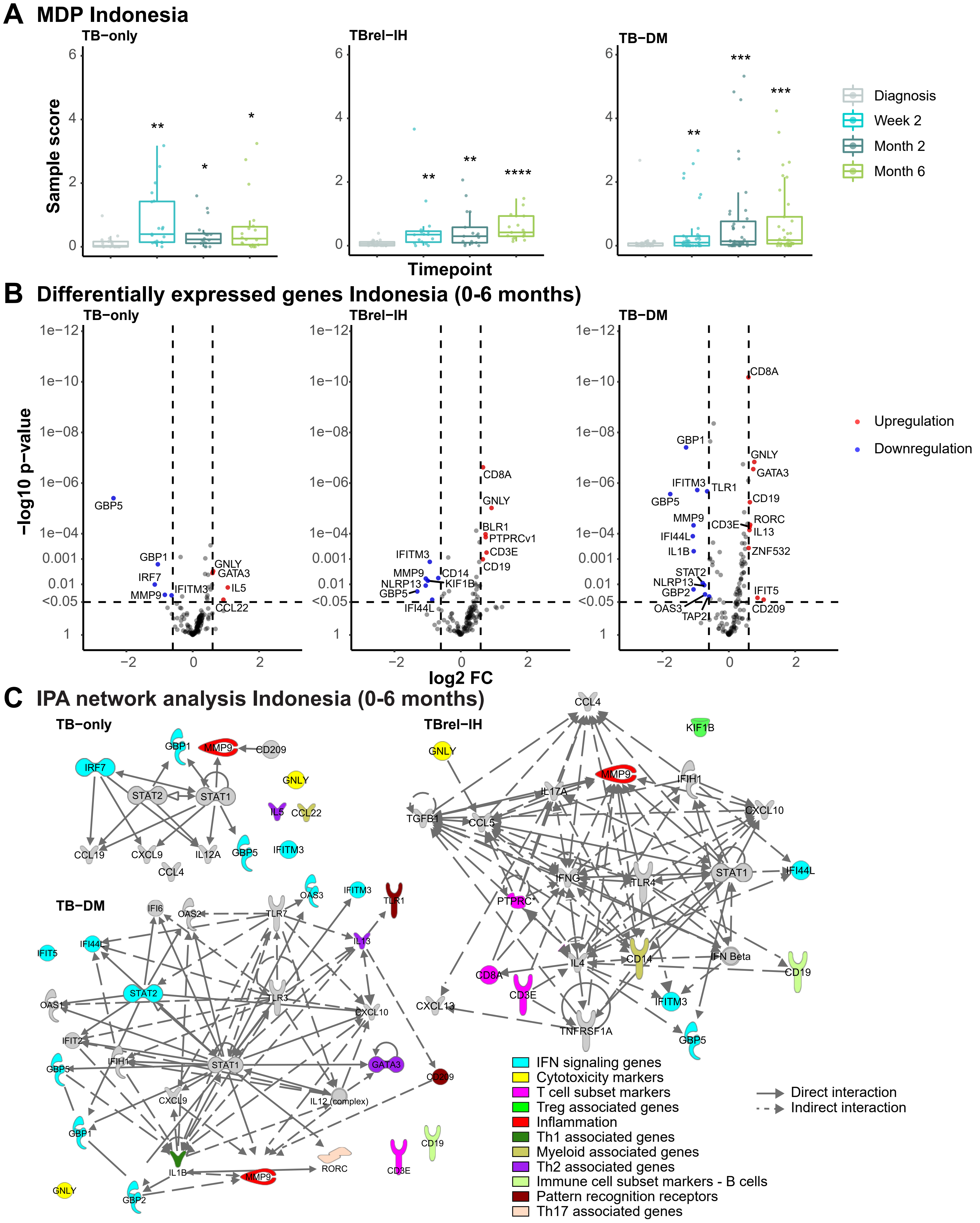


**Supplementary Figure 6. TB treatment response in TB patients is dependent on diabetes/glycaemia status in the Indonesian cohort.** MDP and Differential Expression Analyses were performed on GAPDH-normalized log2-transformed targeted gene expression data of the Indonesian cohort. (A) MDP analyses of the different study groups showing the impact of TB treatment on the overall gene perturbation over time. Samples of patients at diagnosis were used as baseline controls. (B) Volcano plots representing DEGs regulated during treatment of TB patients categorized based on their diabetes/glycaemia status. The y-axis scales of all plots are harmonized per study group. -log10-transformed p-values are plotted against log2 FC. Genes with p <0.05 and log2 FC <-0.6 or >0.6 were labelled as DEGs. (C) IPA interactive network analyses of DEGs regulated during treatment. The various shapes of the nodes represent the functional classes of the gene products. Gene modules are indicated by distinctive colors.

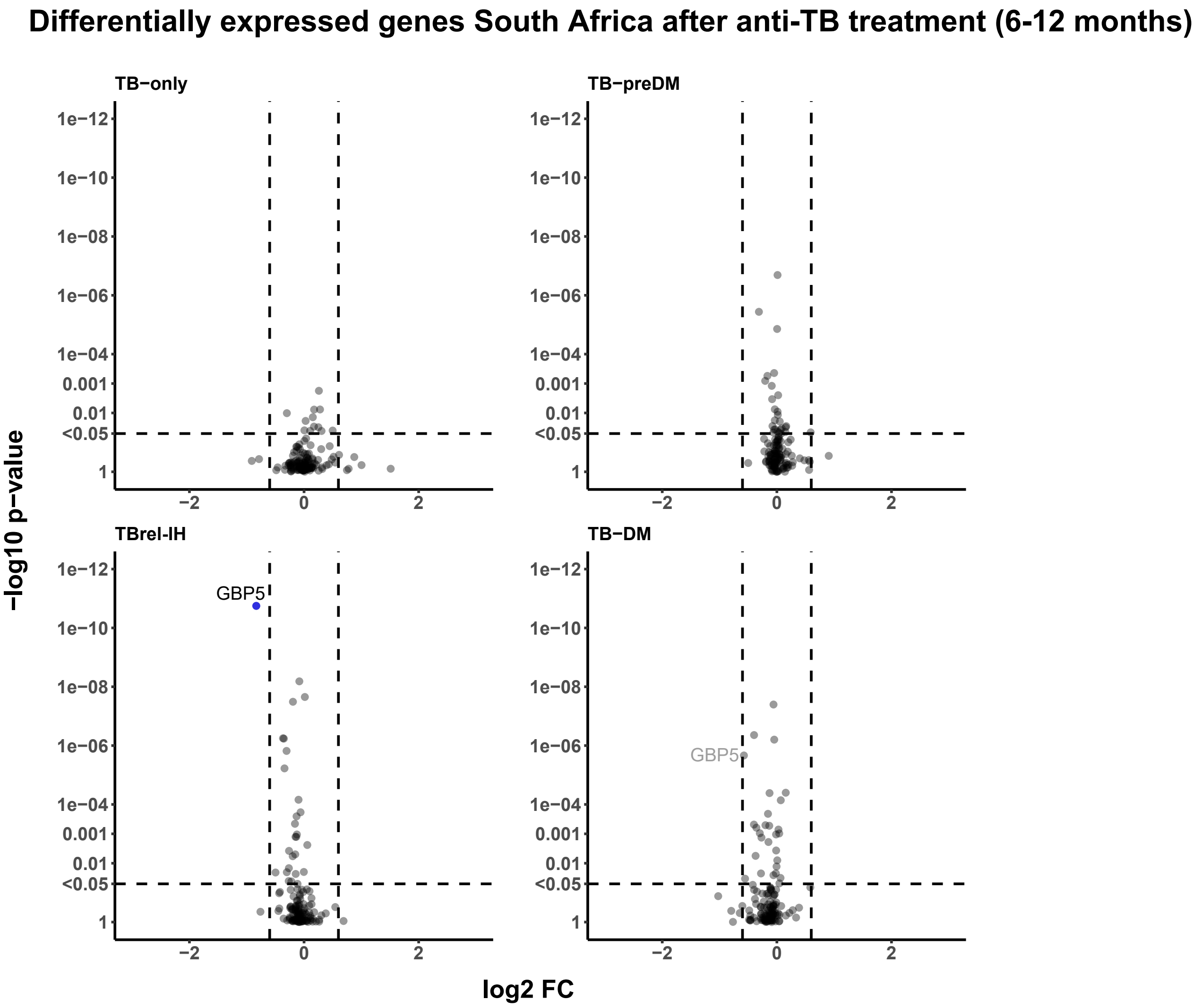


**Supplementary Figure 7. Transcriptional response after anti-TB treatment in the South African cohort.** Volcano plots representing DEGs regulated after TB treatment (6-12 months) of TB patients categorized based on their diabetes/glycaemia status. The y-axis scales of all plots are harmonized per study group. P-values, -log10-transformed for better visualization, are plotted against log2 FC. Genes with p <0.05 and log2 FC <-0.6 or >0.6 were labelled as DEGs.

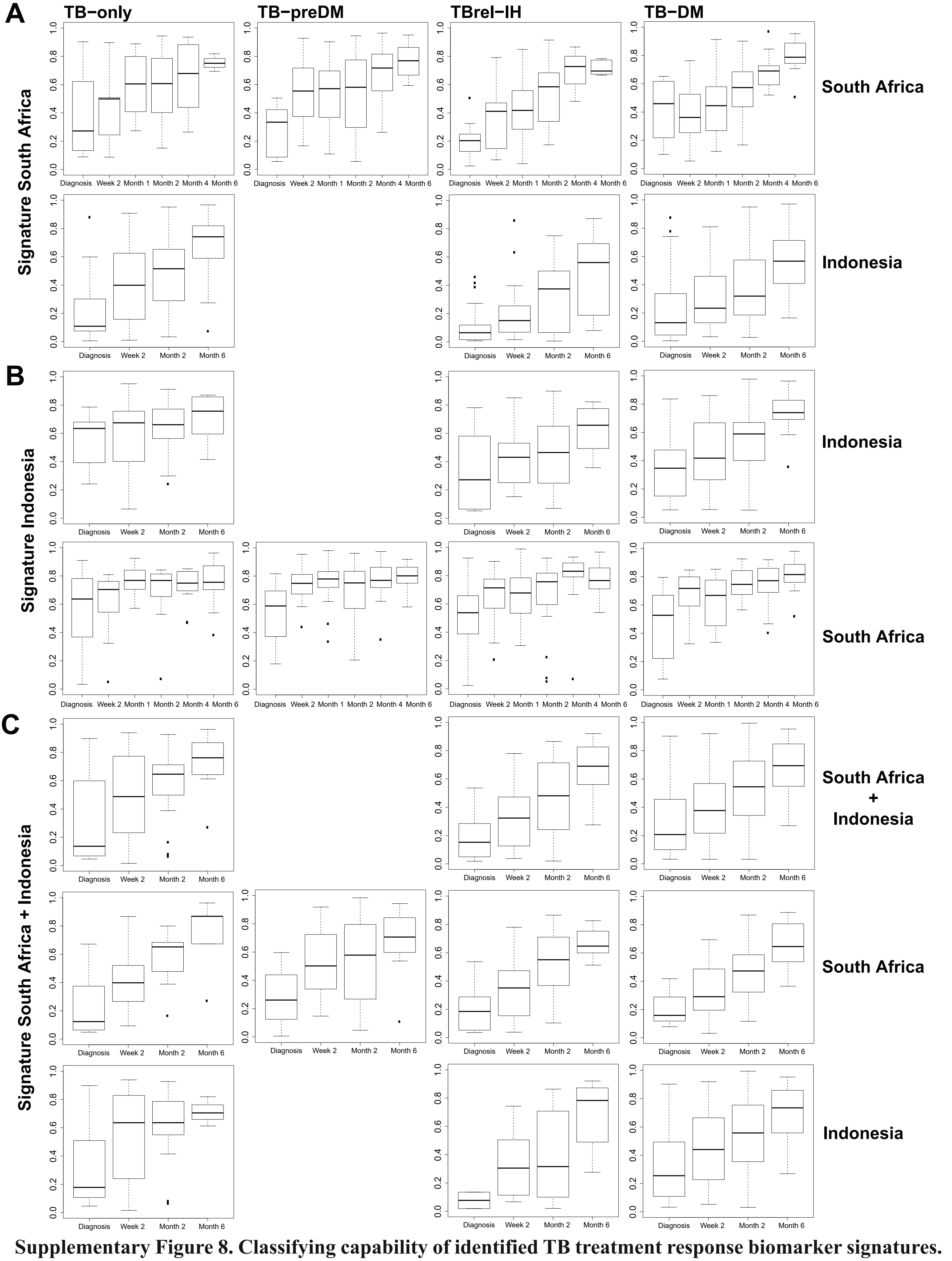


**Supplementary Figure 8. Classifying capability of identified TB treatment response biomarker signatures.** Predicted probability plots showing the accuracy of TB treatment response biomarker signatures identified in (A) South Africa or (B) Indonesia across timepoints in box-and-whiskers plots (5-95 percentiles) either in the cohort in which the Train Test Split (TTS) was performed or in the validation cohort, split into TB study groups based on their diabetes/glycaemia status. (C) Predicted probability plots showing the accuracy of the identified pooled (South Africa and Indonesia) biomarker signature across timepoints in box-and-whiskers plots (5-95 percentiles) either in the pooled cohort in which Train-Test Split (TTS) was performed (left panel) or the single validation cohorts (right panels), split into TB study groups based on their diabetes/glycaemia status.

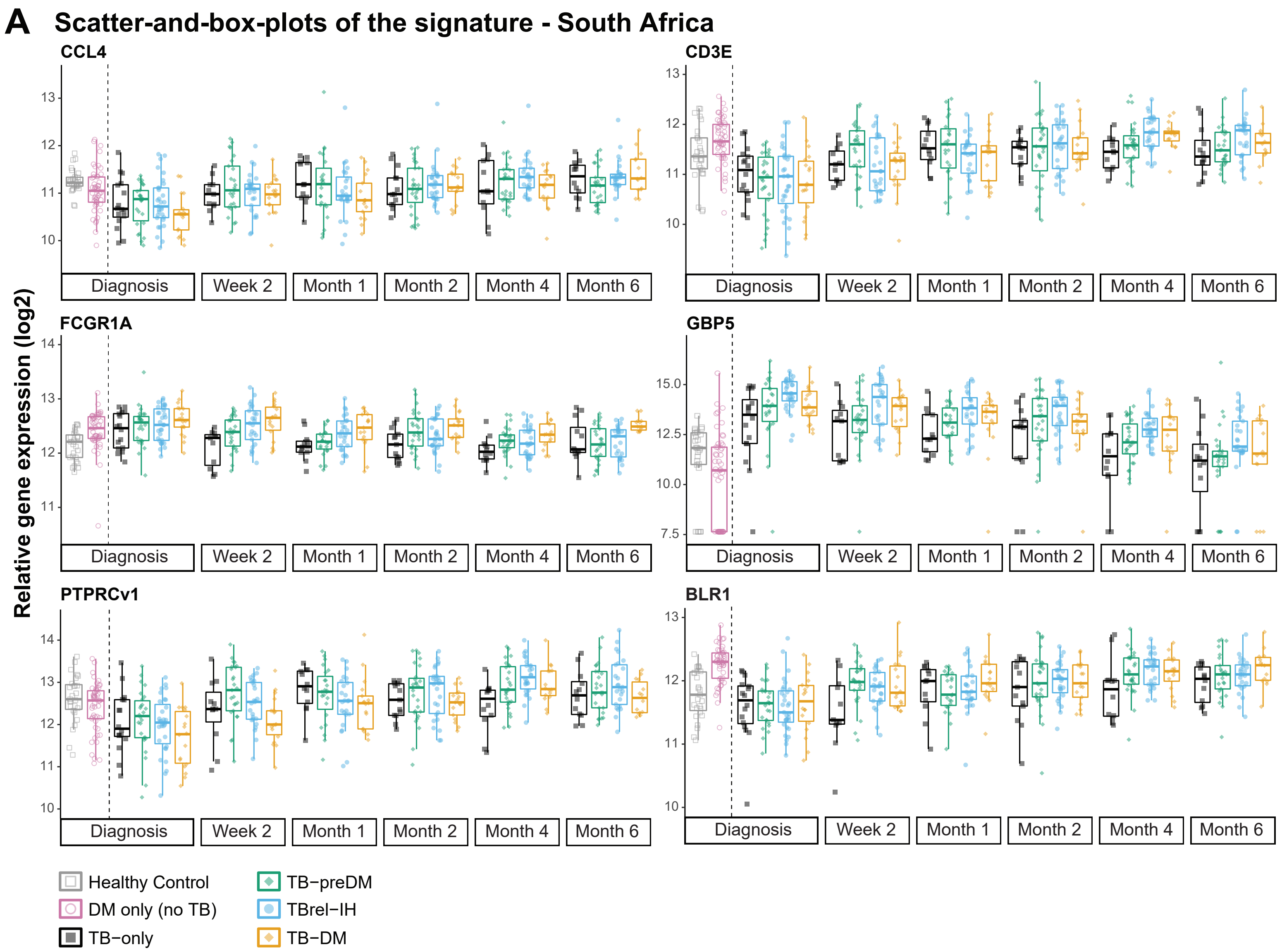


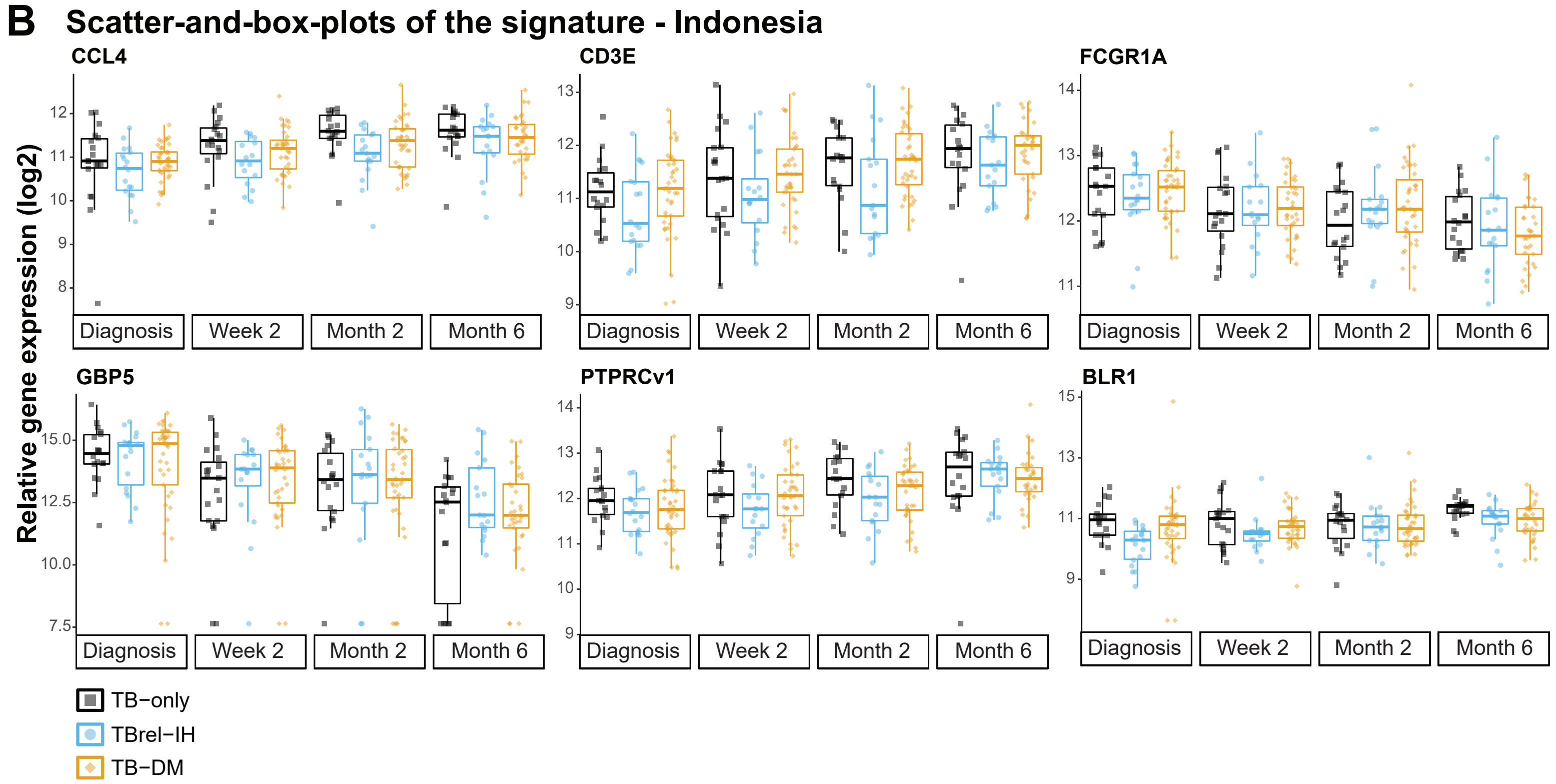


**Supplementary Figure 9. The kinetic profiles of 6 representative genes of the 15-gene TB treatment response signature of the pooled cohort dataset.** Expression kinetics of single genes in the (A) South African cohort or (B) Indonesian cohort at diagnosis and during TB treatment. Box plots depict log2-transformed median gene expression values and the inter quartile range (IQR), while the whiskers represent the data within the Q1-1.5xIQR and Q3+1.5xIQR interval. Outliers are reported as symbols.
