## Supplementary material for "Impaired resolution of blood transcriptomes through tuberculosis treatment with diabetes comorbidity": TANDEM_Full_Consortium

| First Name | Initial | Last Name | email address | Affiliation |
|---|---|---|---|---|
| Hazel | M. | Dockrell | | TB Centre and Department of Immunology and Infection, London School of Hygiene & Tropical Medicine, United Kingdom |
| Jacqueline | M. | Cliff | | TB Centre and Department of Immunology and Infection, London School of Hygiene & Tropical Medicine, United Kingdom |
| Clare | | Eckold | | TB Centre and Department of Immunology and Infection, London School of Hygiene & Tropical Medicine, United Kingdom |
| JiSook | | Lee | | TB Centre and Department of Immunology and Infection, London School of Hygiene & Tropical Medicine, United Kingdom |
| David | A. | Moore | | TB Centre and Department of Clinical Research, London School of Hygiene & Tropical Medicine, United Kingdom |
| Ulla | K. | Griffiths | | Department of Global Health and Development, London School of Hygiene & Tropical Medicine, United Kingdom |
| Yoko | V. | Laurence | | TB Centre and Department of Global Health and Development, London School of Hygiene & Tropical Medicine, United Kingdom |
| Rob | R | Anmontse | | Radboud university medical center, Dpt Clinical Pharmacy, The Netherlands |
| Mihai | | Netea | | Radboud university medical center, Dpt Internal Medicine, The Netherlands |
| Reinout | | van Crevel | | Radboud university medical center, Dpt Internal Medicine, The Netherlands |
| Carolien | | Ruesen | | Radboud university medical center, Dpt Internal Medicine, The Netherlands |
| Ekta | | Lachmandas | | Radboud university medical center, Dpt Internal Medicine, The Netherlands |
| Stefan | H.E. | Kaufmann | | Max Plank Institute for Infection Biology, Berlin, Germany |
| Macarena | | Beigier | | Max Plank Institute for Infection Biology, Berlin, Germany |
| Golinski | | Robert | | Max Plank Institute for Infection Biology, Berlin, Germany |
| Weiner | | January | | Max Plank Institute for Infection Biology, Berlin, Germany |
| Simone | A. | Joosten | | Infectious Diseases, Leiden University Medical Center, The Netherlands |
| Tom | H.M. | Ottenhoff | | Infectious Diseases, Leiden University Medical Center, The Netherlands |
| Frank | | Vrieling | | Infectious Diseases, Leiden University Medical Center, The Netherlands |
| Marielle | C. | Haks | | Infectious Diseases, Leiden University Medical Center, The Netherlands |
| Gerhard | | Walzl | | SA MRC Centre for TB Research, DST/NRF Centre of Excellence for Biomedical Tuberculosis Research, Faculty of Medicine and Health Sciences, Stellenbosch University, Cape Town, South Africa |
| Katharina | | Ronacher | | SA MRC Centre for TB Research, DST/NRF Centre of Excellence for Biomedical Tuberculosis Research, Faculty of Medicine and Health Sciences, Stellenbosch University, Cape Town, South Africa; The University of Queensland, Translational Research Institute, Brisbane, Australia |
| Stephanus | T. | Malherbe | | SA MRC Centre for TB Research, DST/NRF Centre of Excellence for Biomedical Tuberculosis Research, Faculty of Medicine and Health Sciences, Stellenbosch University, Cape Town, South Africa |
| Léanie | | Kleynhans | | SA MRC Centre for TB Research, DST/NRF Centre of Excellence for Biomedical Tuberculosis Research, Faculty of Medicine and Health Sciences, Stellenbosch University, Cape Town, South Africa |
| Bronwyn | | Smith | | SA MRC Centre for TB Research, DST/NRF Centre of Excellence for Biomedical Tuberculosis Research, Faculty of Medicine and Health Sciences, Stellenbosch University, Cape Town, South Africa |
| Kim | | Stanley | | SA MRC Centre for TB Research, DST/NRF Centre of Excellence for Biomedical Tuberculosis Research, Faculty of Medicine and Health Sciences, Stellenbosch University, Cape Town, South Africa |
| Gian | D. | van der Spuy | | SA MRC Centre for TB Research, DST/NRF Centre of Excellence for Biomedical Tuberculosis Research, Faculty of Medicine and Health Sciences, Stellenbosch University, Cape Town, South Africa |
| André | G. | Loxton | | SA MRC Centre for TB Research, DST/NRF Centre of Excellence for Biomedical Tuberculosis Research, Faculty of Medicine and Health Sciences, Stellenbosch University, Cape Town, South Africa |
| Novel | N. | Chegou | | SA MRC Centre for TB Research, DST/NRF Centre of Excellence for Biomedical Tuberculosis Research, Faculty of Medicine and Health Sciences, Stellenbosch University, Cape Town, South Africa |
| Marika | | Bosman | | SA MRC Centre for TB Research, DST/NRF Centre of Excellence for Biomedical Tuberculosis Research, Faculty of Medicine and Health Sciences, Stellenbosch University, Cape Town, South Africa |
| Leani | | Thiart | | SA MRC Centre for TB Research, DST/NRF Centre of Excellence for Biomedical Tuberculosis Research, Faculty of Medicine and Health Sciences, Stellenbosch University, Cape Town, South Africa |
| Chandré | | Wagman | | SA MRC Centre for TB Research, DST/NRF Centre of Excellence for Biomedical Tuberculosis Research, Faculty of Medicine and Health Sciences, Stellenbosch University, Cape Town, South Africa |
| Happy | | Tshivhula | | SA MRC Centre for TB Research, DST/NRF Centre of Excellence for Biomedical Tuberculosis Research, Faculty of Medicine and Health Sciences, Stellenbosch University, Cape Town, South Africa |
| Mosa | | Selamolela | | SA MRC Centre for TB Research, DST/NRF Centre of Excellence for Biomedical Tuberculosis Research, Faculty of Medicine and Health Sciences, Stellenbosch University, Cape Town, South Africa |
| Nicole | | Prins | | SA MRC Centre for TB Research, DST/NRF Centre of Excellence for Biomedical Tuberculosis Research, Faculty of Medicine and Health Sciences, Stellenbosch University, Cape Town, South Africa |
| Willem | J. | du Plessis | | SA MRC Centre for TB Research, DST/NRF Centre of Excellence for Biomedical Tuberculosis Research, Faculty of Medicine and Health Sciences, Stellenbosch University, Cape Town, South Africa |
| Ilana | C. | van Rensburg | | SA MRC Centre for TB Research, DST/NRF Centre of Excellence for Biomedical Tuberculosis Research, Faculty of Medicine and Health Sciences, Stellenbosch University, Cape Town, South Africa |
| Lorinda | | du Toit | | SA MRC Centre for TB Research, DST/NRF Centre of Excellence for Biomedical Tuberculosis Research, Faculty of Medicine and Health Sciences, Stellenbosch University, Cape Town, South Africa |
| Julia | A | Critchley | | Population Health Research Institute, St George's, University of London, United Kingdom |
| Sarah | R | Kerry | | Population Health Research Institute, St George's, University of London, United Kingdom |
| Fiona | | Pearson | | Population Health Research Institute, St George's, University of London, United Kingdom |
| Daniel | | Grint | | Population Health Research Institute, St George's, University of London, United Kingdom |
| MIHAI | | IOANA | | Human Genomics Laboratory, Universitatea de Medicină si Farmacie din Craiova, Romania; Dolj Regional Centre of Medical Genetics, Spitalul Clinic Județean de Urgență Craiova, Romania |
| MIRCEA NICOLAE | | PANDURU | | Human Genomics Laboratory, Universitatea de Medicina si Farmacie din Craiova, Romania |
| ANCA | L | RIZA | | Human Genomics Laboratory, Universitatea de Medicină și Farmacie din Craiova, Romania; Department of Internal Medicine and Radboud Center for Infectious Diseases, Radboud University Medical Center, Nijmegen, The Netherlands |
| RAMONA | | CIOBOATA | | Internal Medicine - Pulmunology Dept., Universitatea de Medicină și Farmacie din Craiova, Romania; Pneumophtisiology Clinic, Spitalul Clinic de Boli Infecțioase și Pneumoftiziologie "Victor Babeş" Craiova, Romania |
| MIHAELA | O | DUDAU | | Pneumophtisiology Clinic, Spitalul Clinic de Boli Infecțioase și Pneumoftiziologie "Victor Babeş" Craiova, Romania |
| FLOAREA | M | NITU | | Internal Medicine - Phtisiology Dept., Universitatea de Medicină și Farmacie din Craiova, Romania; Pneumophtisiology Clinic, Spitalul Clinic de Boli Infecțioase și Pneumoftiziologie "Victor Babeş" Craiova, Romania |
| ILEANA | C | BAZAVAN | | Internal Medicine - Phtisiology Dept., Universitatea de Medicină și Farmacie din Craiova, Romania; Pneumophtisiology Clinic, Spitalul Clinic de Boli Infecțioase și Pneumoftiziologie "Victor Babeş" Craiova, Romania |
| MIHAI | | OLTEANU | | Internal Medicine - Phtisiology Dept., Universitatea de Medicină și Farmacie din Craiova, Romania; Pneumophtisiology Clinic, Spitalul Clinic de Boli Infecțioase și Pneumoftiziologie "Victor Babeş" Craiova, Romania |
| CORNELIA | D. | EDITOIU | | TB laboratory, Spitalul Clinic de Boli Infecțioase și Pneumoftiziologie "Victor Babeş" Craiova, Romania |
| ADRIANA | | FLORESCU | | TB laboratory, Spitalul Clinic de Boli Infecțioase și Pneumoftiziologie "Victor Babeş" Craiova, Romania |
| MARIUS | S | CIONTEA | | Pulmunology Clinic, Spitalul de Pneumoftiziologie "Tudor Vladimirescu", com. Runcu, Gorj, Romania |
| IULIA | D. | CAPITANESCU | | Pulmunology Clinic, Spitalul de Pneumoftiziologie "Tudor Vladimirescu", com. Runcu, Gorj, Romania |
| MARIAN | | OLARU | | Pulmunology Clinic, Spitalul de Pneumoftiziologie "Tudor Vladimirescu", com. Runcu, Gorj, Romania |
| TIBERIU | | TATARU | | Pulmunology Clinic, Spitalul de Pneumoftiziologie "Tudor Vladimirescu", com. Runcu, Gorj, Romania |
| MARIA | D. | PAPURICA | | TB laboratory, Spitalul de Pneumoftiziologie "Tudor Vladimirescu", com. Runcu, Gorj, Romania |
| ILEANA | | VALUTANU | | TB laboratory, Spitalul de Pneumoftiziologie "Tudor Vladimirescu", com. Runcu, Gorj, Romania |
| VASILICA | | DUBREU | | TB laboratory, Spitalul de Pneumoftiziologie "Tudor Vladimirescu", com. Runcu, Gorj, Romania |

| | | | | |
|---|---|---|---|---|
| LIVIU | | STAMATOIU | | TB laboratory, Spitalul de Pneumoftiziologie "Tudor Vladimirescu", com. Runcu, Gorj, Romania |
| CREOLA | | ENOIU | | TB laboratory, Spitalul Județean de Urgență Targu-Jiu, Romania |
| MARIA | | MOTA | | Nutition and Metabolic Disorders Dept, Universitatea de Medicină şi Farmacie din Craiova, Romania; Diabetes, nutrition and metabolic disorders clinic, Spitalul Clinic Județean de Urgență Craiova, Romania |
| SIMONA-GEORGIANA | | POPA | | Nutition and Metabolic Disorders Dept, Universitatea de Medicină şi Farmacie din Craiova, Romania; Diabetes, nutrition and metabolic disorders clinic, Spitalul Clinic Județean de Urgență Craiova, Romania |
| ADELA | G. | FIRANESCU | | Diabetes, Nutrition and Metabolic Disorders Clinic, Spitalul Clinic Județean de Urgență Craiova, Romania |
| ADINA | | POPA | | Diabetes, Nutrition and Metabolic Disorders Clinic, Spitalul Clinic Județean de Urgență Craiova, Romania |
| IOANA | A | GHEONEA | | Radiology and Imaging Dept., Universitatea de Medicină şi Farmacie din Craiova, Romania; Radiology and Medical Imaging Laboratory, Spitalul Clinic Județean de Urgență Craiova, Romania |
| STEFANIA | | BICUTI | | Radiology and Medical Imaging Laboratory, Spitalul Clinic Județean de Urgență Craiova, Romania |
| ALINA | | LEPADAT | | Radiology and Medical Imaging Laboratory, Spitalul Clinic Județean de Urgență Craiova, Romania |
| IONELA MIHAELA | | VLADU | | Nutition and Metabolic Disorders Dept, Universitatea de Medicină şi Farmacie din Craiova, Romania; Diabetes, Nutrition and Metabolic Disorders Comp., Internal Medicine Dept., Spitalul Clinic Municipal Filantropia Craiova, Romania |
| DIANA | | CLENCIU | | Diabetes, Nutrition and Metabolic Disorders Comp., Internal Medicine Dept., Spitalul Clinic Municipal Filantropia Craiova, Romania |
| MIHAELA | L | BICU | | Diabetes, Nutrition and Metabolic Disorders Comp., Internal Medicine Dept., Spitalul Clinic Municipal Filantropia Craiova, Romania |
| COSTIN | | STREBA | | Internal Medicine - Pulmunology Dept., Universitatea de Medicină şi Farmacie din Craiova, Romania |
| ALIN | D. | DEMETRIAN | | Thoracic Surgery Dept., Universitatea de Medicină şi Farmacie din Craiova, Romania; Thoracic Surgery Clinic, Spitalul Clinic Județean de Urgență Craiova, Romania |
| MARIUS | | CIUREA | | Thoracic Surgery Dept., Universitatea de Medicină şi Farmacie din Craiova, Romania; Thoracic Surgery Clinic, Spitalul Clinic Județean de Urgență Craiova, Romania |
| ALINA | | CIMPOERU | | Human Genomics Laboratory, Universitatea de Medicină si Farmacie din Craiova, Romania |
| ADELA | | CIOCOIU | | Human Genomics Laboratory, Universitatea de Medicină si Farmacie din Craiova, Romania; Dolj Regional Centre of Medical Genetics, Spitalul Clinic Județean de Urgență Craiova, Romania |
| STEFANIA | C | DOROBANTU | | Human Genomics Laboratory, Universitatea de Medicină si Farmacie din Craiova, Romania |
| RAZVAN | M | PLESEA | | Human Genomics Laboratory, Universitatea de Medicină si Farmacie din Craiova, Romania; Dolj Regional Centre of Medical Genetics, Spitalul Clinic Județean de Urgență Craiova, Romania |
| ELENA-LEOCARDIA | | POPESCU | | Human Genomics Laboratory, Universitatea de Medicină si Farmacie din Craiova, Romania; Dolj Regional Centre of Medical Genetics, Spitalul Clinic Județean de Urgență Craiova, Romania |
| MIHAI | G. | CUCU | | Human Genomics Laboratory, Universitatea de Medicină si Farmacie din Craiova, Romania; Dolj Regional Centre of Medical Genetics, Spitalul Clinic Județean de Urgență Craiova, Romania |
| IOANA | | STREATA | | Human Genomics Laboratory, Universitatea de Medicină si Farmacie din Craiova, Romania; Dolj Regional Centre of Medical Genetics, Spitalul Clinic Județean de Urgență Craiova, Romania |
| FLORIN | | BURADA | | Human Genomics Laboratory, Universitatea de Medicină si Farmacie din Craiova, Romania; Dolj Regional Centre of Medical Genetics, Spitalul Clinic Județean de Urgență Craiova, Romania |
| SIMONA | | SERBAN-SOSOI | | Human Genomics Laboratory, Universitatea de Medicină si Farmacie din Craiova, Romania |
| ELENA | R | NICOLI | | Human Genomics Laboratory, Universitatea de Medicină si Farmacie din Craiova, Romania |
| Susan | M | McAllister | | Centre for International Health, University of Otago, New Zealand |
| Philip | C | Hill | | Centre for International Health, University of Otago, New Zealand |
| Ajesha | E | Verrall | | Centre for International Health, University of Otago, New Zealand |
| Vinod | | Kumar | | Department of Genetics, University of Groningen, The Netherlands |
| Cisca | | Wijmenga | | Department of Genetics, University of Groningen, The Netherlands |
| Cesar | | Ugarte-Gil | | Facultad de Medicina Alberto Hurtado, Universidad Peruana Cayetano Heredia, Lima, Perú |
| Jorge | | Coronel | | Laboratorio de Investigación de Enfermedades Infecciosas, Universidad Peruana Cayetano Heredia, Lima, Perú |
| Sonia | | Lopez | | Laboratorio de Investigación de Enfermedades Infecciosas, Universidad Peruana Cayetano Heredia, Lima, Perú |
| Ruth | | Limascca | | Laboratorio de Investigación de Enfermedades Infecciosas, Universidad Peruana Cayetano Heredia, Lima, Perú |
| Katherine | | Villaizan | | Laboratorio de Investigación de Enfermedades Infecciosas, Universidad Peruana Cayetano Heredia, Lima, Perú |
| Beatriz | | Castro | | Laboratorio de Investigación de Enfermedades Infecciosas, Universidad Peruana Cayetano Heredia, Lima, Perú |
| Jhomelin | | Flores | | Laboratorio de Investigación de Enfermedades Infecciosas, Universidad Peruana Cayetano Heredia, Lima, Perú |
| Walter | | Solano | | Laboratorio de Investigación de Enfermedades Infecciosas, Universidad Peruana Cayetano Heredia, Lima, Perú |
| Bachti | BA | Alisjahbana | | Internal Medicine, Infectious Disease, Universitas Padjadjaran, Bandung, Indonesia |
| Rovina | NR | Ruslami | | Pharmacology and Therapy, Universitas Padjadjaran, Bandung, Indonesia |
| Nanny NM | NNS | Soetedjo | | Internal Medicine, Endocrinology, Universitas Padjadjaran, Bandung, Indonesia |
| Prayudi | PP | Santoso | | Internal Medicine, Pulmonology, Universitas Padjadjaran, Bandung, Indonesia |
| Lidya | LC | Chaidir | | TB-HIV Research Center, Universitas Padjadjaran, Bandung, Indonesia |
| Raspati C | RCK | Koesoemadina | | Microbiology and Parasitology, Universitas Padjadjaran, Bandung, Indonesia |
| Nopi | NS | Susilawati | | TB-HIV Research Center, Universitas Padjadjaran, Bandung, Indonesia |
| Jessi | JA | Annisa | | TB-HIV Research Center, Universitas Padjadjaran, Bandung, Indonesia |
| Resvi | RL | Livia | | TB-HIV Research Center, Universitas Padjadjaran, Bandung, Indonesia |
| Vycke | VY | Yunivita | | Pharmacology and Therapy, Universitas Padjadjaran, Bandung, Indonesia |
| Arto Y | AYS | Soeroto | | Internal Medicine, Pulmonology, Universitas Padjadjaran, Bandung, Indonesia |
| Hikmat | HP | Permana | | Internal Medicine, Endocrinology, Universitas Padjadjaran, Bandung, Indonesia |
| Sofia | SI | Imaculata | | TB-HIV Research Center, Universitas Padjadjaran, Bandung, Indonesia |
| Yuanita | YG | Gunawan | | TB-HIV Research Center, Universitas Padjadjaran, Bandung, Indonesia |
| Nury Fitria | NFD | Dewi | | TB-HIV Research Center, Universitas Padjadjaran, Bandung, Indonesia |
| Lika Apriani | LA | Apriani | | Public Health, Universitas Padjadjaran, Bandung, Indonesia |
